# Could nutrition modulate COVID-19 susceptibility and severity of disease? A systematic review

**DOI:** 10.1101/2020.10.19.20214395

**Authors:** Philip T. James, Zakari Ali, Andrew E. Armitage, Ana Bonell, Carla Cerami, Hal Drakesmith, Modou Jobe, Kerry S. Jones, Zara Liew, Sophie E. Moore, Fernanda Morales-Berstein, Helen M. Nabwera, Behzad Nadjm, Sant-Rayn Pasricha, Pauline Scheelbeek, Matt J. Silver, Megan R. Teh, Andrew M. Prentice

## Abstract

**Background:** Many nutrients have powerful immunomodulatory actions with the potential to alter susceptibility to COVID-19 infection, progression to symptoms, likelihood of severe disease and survival. The pandemic has fostered many nutrition-related theories, sometimes backed by a biased interpretation of evidence.

**Objectives:** To provide a systematic review of the latest evidence on how malnutrition across all its forms (under- and over-nutrition and micronutrient status) may influence both susceptibility to, and progression and severity of, COVID-19.

**Methods:** We synthesised information on 13 nutrition-related components and their potential interactions with COVID-19: overweight, obesity and diabetes; protein-energy malnutrition; anaemia; vitamins A, C, D, and E; poly-unsaturated fatty acids; iron; selenium; zinc; anti-oxidants, and nutritional support. For each section we provide: a) a landscape review of pertinent material; b) a systematic search of the literature in PubMed and EMBASE databases, including a systematic search of a wide range of pre-print servers; and c) a screen of six clinical trial registries. Two reviewers were assigned per section for data extraction. All original research was considered, without restriction to study design, and included if it covered: 1) SARS-CoV-2, MERS-CoV or SARS-CoV viruses and 2) disease susceptibility or 3) disease progression, and 4) the nutritional component of interest. Searches took place between 16^th^ May and 11^th^ August, 2020. PROSPERO registration CRD42020186194.

**Results:** Across the 13 searches, a total of 2732 articles from PubMed and EMBASE, 4164 articles from the pre-print servers, and 433 trials were returned. A total of 288 published articles and 278 pre-print articles were taken to full text screening. In the final narrative synthesis, we cover 22 published articles, 39 pre-print articles and 79 trials. The review highlights a range of mechanistic and observational evidence to highlight the role nutrition can play in susceptibility and progression of COVID-19. However, to date, there is limited evidence that high-dose supplements of micronutrients will either prevent severe disease or speed up recovery, although results of clinical trials are eagerly awaited.

**Conclusions:** To date there is no conclusive evidence supporting adoption of novel nutritional therapies. However, given the known impacts of all forms of malnutrition on the immune system, public health strategies to reduce micronutrient deficiencies and undernutrition remain of critical importance. There is strong evidence that prevention of obesity, and its consequent type-2 diabetes, will reduce the risk of serious COVID-19 outcomes.

## 1. Introduction

The astonishing spread of severe acute respiratory syndrome coronavirus-2 (SARS-CoV-2, Box 1) since late 2019 has resulted in a global pandemic of the disease COVID-19. Alongside the worldwide effort to deliver a vaccine, there has been a surge of interest in the epidemiological factors that underlie susceptibility to COVID-19, and its progression, in an attempt to explore the most effective preventative and curative options^1–4^. Potential interactions between nutritional status and immune function have been widely documented^5–7^. As the pandemic unfolds, it exacerbates the risk factors for malnutrition in all its forms^8,9^. Disruption to agricultural production, market linkages and seasonal labour movements contribute to food price increases^10,11^, making nutritious food even more expensive for those most at risk of micronutrient deficiencies and undernutrition. Cancelled and delayed nutrition counselling, micronutrient distributions, vaccine rounds and school meal programmes accentuate the vulnerability^12–14^. Lockdown measures in many countries have increased physical and psychological barriers to healthy eating and exercising, creating an obesogenic environment for many^15,16^.

Understanding the relationship between nutritional status and risk of COVID-19 is therefore of critical importance to generate evidence-based recommendations. There may be a potential for nutritional interventions to reduce an individual’s susceptibility to infection, progression to symptoms and likelihood of severe disease (including the use of high- or very-high-dose supplements enterally or intravenously as nutraceuticals).

However, nutrition information has long been miscommunicated to the public^17–19^, and nutrition-related myths on COVID-19 protection and treatment are widely prevalent in this pandemic^20^. To this end we have conducted a comprehensive systematic review of journal articles, pre-prints and clinical trial registries to provide a robust evidence base of what is currently known and what gaps remain.

### Box 1

**Coronaviruses and COVID-19**

Coronaviruses consist of a small single-stranded RNA, belong to the Coronaviridae family. There are four sub-groups (α, β, γ and δ), of which the α- and β-coronaviruses are known to infect humans from zoonotic origins^21,22^. Coronavirus infection rates can vary seasonally due in part to the underlying epidemiology of susceptible host availability^23^.

The pathogenicity of coronavirus infections in humans became apparent with the severe acute respiratory syndrome coronavirus (SARS-CoV) causing an outbreak of SARS in 2002-3, originating in Guangdong, China^24^. A decade later, the Middle East respiratory syndrome coronavirus (MERS-CoV) was first detected in 2012 in Saudi Arabia^25^. COVID-19, the disease caused by the severe acute respiratory syndrome coronavirus-2 (SARS-CoV-2), originated in Wuhan, China in late 2019. It was declared a global pandemic by the World Health Organisation on 11 March 2020. SARS-CoV-2 is a β-coronavirus and, as with SARS-CoV and MERS-CoV, can cause dysregulation of the pulmonary vasculature, microthromboembolisms, pneumonia and may progress to acute respiratory distress syndrome (ARDS), multi-system organ failure and death^26–28^. SARS-CoV-2 invades type II alveolar epithelial cells, accessing cellular machinery through the binding of its spike protein to Angiotensin-converting enzyme 2 (ACE2), which is highly expressed in the lungs and heart^28^. To date, SARS-CoV-2 exhibits higher transmissibility but lower mortality than SARS-CoV and MERS-CoV^29^.

## 2. Methods

This review considers how malnutrition across all its forms (undernutrition, micronutrient deficiencies and overnutrition) may influence both susceptibility to, and progression of, COVID-19. We synthesised information on 13 nutrition-related components and their potential interactions with COVID-19: overweight, obesity and diabetes; protein-energy malnutrition; anaemia; vitamins A, C, D, and E; poly-unsaturated fatty acids; iron; selenium; zinc; anti-oxidants, and nutritional support. We published our strategy on the PROSPERO database, reference CRD42020186194.

### Search Strategy

We adopted three key approaches for compiling information for each of the 13 sections listed above:

a. A landscape review of pertinent material. This section is non-systematic, and covers a brief description of the nutrient/condition vis-à-vis infection and immunity, evidence of any role in viral infections, possible mechanisms, and possible utility in treatment.
b. A systematic search of the literature in PubMed and EMBASE databases, and including a systematic search of a wide range of pre-print servers (listed in Supplementary Material 1).
c. A screen of six clinical trial registries, listed in Supplementary Material 1.

For the PubMed and EMBASE database searches a search string was designed to encompass terms related to 1) SARS-CoV-2, MERS-CoV or SARS-CoV viruses, 2) disease susceptibility, 3) disease progression and 4) the nutritional component of interest. The search string was then built combining the terms for 1 AND (2 OR 3) AND 4. The search string corresponding to components 1-3 was kept consistent between all sections, with component 4 being adapted to the relevant exposure of interest. The clinical trial registry and pre-print server searches were restricted to COVID-19. Full search string terms for the PubMed, EMBASE, pre-print server and clinical trial registry searches are provided in Supplementary Material 2.

In the landscape reviews we summarised the insights learnt from other viral diseases where relevant, and included other coronaviruses (MERS-CoV and SARS-CoV) in the systematic searches. From the outset we acknowledge that COVID-19 is behaving differently to other viral diseases, and therefore cautiously extrapolate risk throughout the review.

### Inclusion and exclusion criteria

We considered all populations of any sex, age, or nutritional status, with no specific geographic boundaries. We restricted the systematic searches to human populations and studies in English. All original research was considered, without restriction to study design. Systematic reviews were included to search bibliographies. We excluded comments, letters, opinions and non-systematic reviews.

### Outcomes

Main outcomes for disease susceptibility were related to key concepts such as immunosuppression, inflammation, lymphocyte regulation, oxidative stress and all forms of immune dysfunction. Main outcomes for disease progression related to viral load, viral replication, viral mutation and transmission, worsening of respiratory tract and gastrointestinal infections, multiple organ failure, and other pathological features on disease progression to death. As the potential role of nutrition in disease susceptibility and progression is broad, we did not pre-specify the measures of effect to consider. Instead, we report the measures of effect that the authors have used in the eligible studies.

### Screening and selection

A lead and co-author were assigned to each of the 13 nutrition-related sections of the review. The two researchers then performed the PubMed and EMBASE searches for their section. After abstract screening, full texts were retrieved for the potentially eligible studies. The lead author then reviewed these studies and used a standardised template to extract data on the eligible studies.

A team of two researchers searched and abstract-screened all the pre-print servers listed in Appendix 1 for all 13 sections. They exported potentially eligible matches to the lead author of the relevant section for full screen. One researcher searched all the clinical trial registries for the 13 sections. Details of the potentially eligible clinical trials were sent to the lead author for review and data extraction. Searches took place between 16^th^ May and 11^th^ August, 2020. Full details of the search dates by section can be found in Supplementary Material 3.

Due to the expected heterogeneity of study types, exposures and outcomes, we did not undertake a formal risk of bias assessment for each included study.

### Data synthesis

We were guided by the Synthesis Without Meta-analysis (SWiM) reporting guidelines for systematic reviews^30^. Due to the heterogeneity of outcomes related to disease susceptibility and progression we did not attempt to transform results into a standardised metric. For each section of the review we summarised the effect sizes as reported by the authors in the included studies.

## 3. Results

Figure 1 provides the overall flow chart summary of all articles retrieved and included in the narrative synthesis. The detailed flow chart breakdowns per section are given in Supplementary Material 3. Across the 13 searches, a total of 2732 hits from PubMed and EMBASE were returned. After removal of 661 duplicates, 2071 were taken to title/abstract screen and 1783 were deemed ineligible at this stage. A total of 288 articles were taken to full text screen and 266 were further excluded. The remaining 22 articles were included in the narrative synthesis and further information captured in Supplementary Material 5.

**Figure 1:**
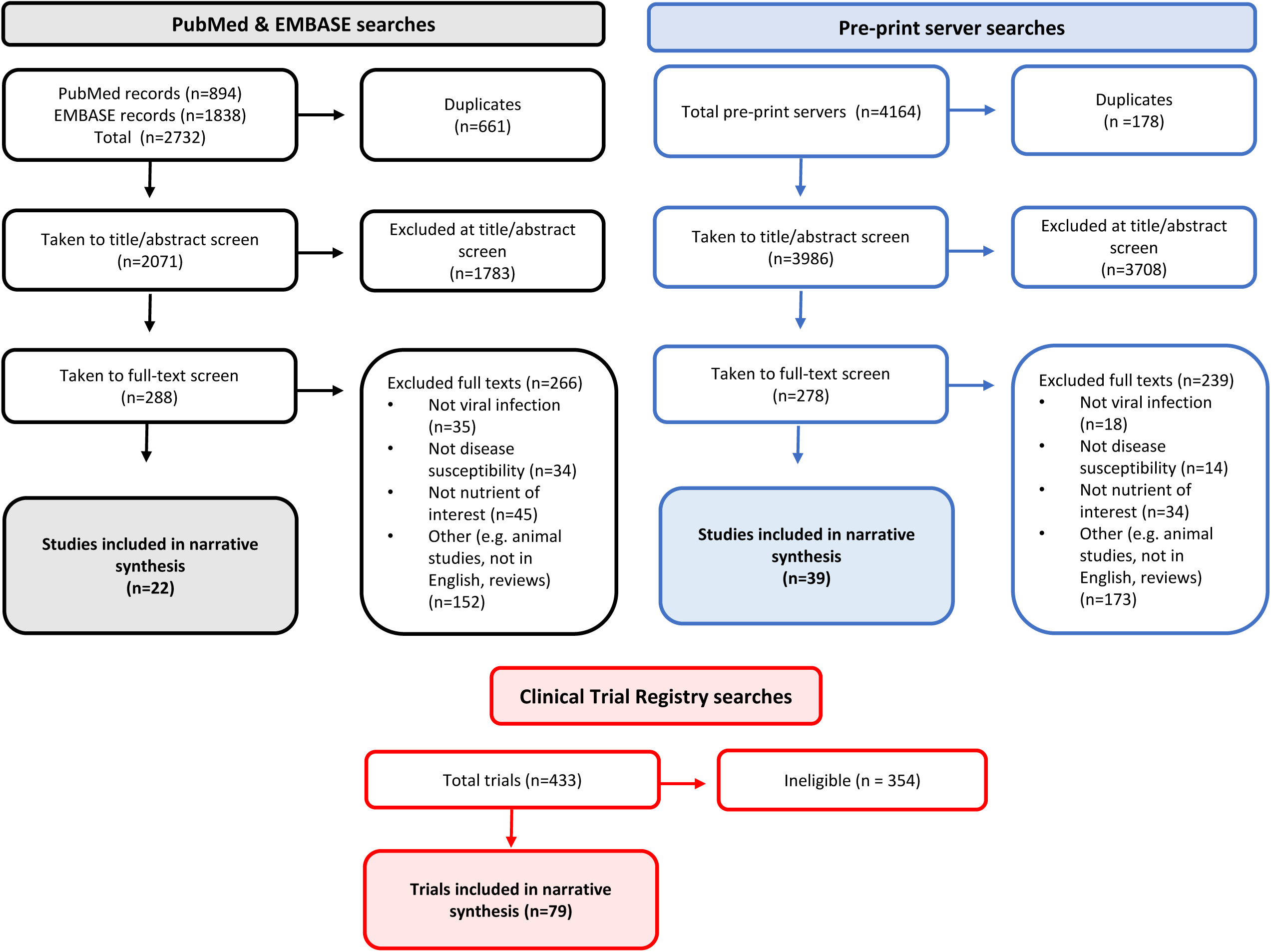
Overview flowchart of articles considered in narrative synthesis.

A total of 4164 hits from across the pre-print servers were returned. After removal of 178 duplicates, 3986 were taken to title/abstract screen and 3708 were ineligible. 278 articles were taken to full text screen and 239 were excluded. The remaining 39 articles were included in the narrative synthesis and Supplementary Material 5.

From the clinical registry searches 433 trials were returned and 354 were ineligible. 79 trials were therefore included in the narrative synthesis and also detailed in Supplementary Material 4.

## 4. Protein Energy Malnutrition

### Landscape review

Protein-energy malnutrition (PEM), also called protein energy undernutrition or simply ‘undernutrition’, is a state of nutritional insufficiency attributable to inadequate energy and/or protein intake, and is often associated with multiple micronutrient deficiencies^31^. According to the 2020 Global Nutrition Report, an estimated 820 million people worldwide (11% of global population) are hungry or undernourished, and the majority are found in low-and-middle income countries (LMICs)^32^.

Globally PEM affects at least 1 in 5 children under 5 years with the greatest burden in LMICs, predominantly those in sub-Saharan African and South Asia^32^. It manifests as stunting (weight-for-age z-scores <-2, compared to the WHO Growth Reference Standards^33^), underweight (including low birth weight, weight-for-age z-scores <-2), and acute malnutrition (kwashiorkor or wasting, defined as weight-for-height/length <-2 z-scores). The severe form of the latter, severe acute malnutrition (SAM), is associated with up to 50% mortality among children admitted to hospital^34^. In 2019, 49.5 million (7.3%) children aged under five years were wasted and 149 million (22%) were stunted globally^32^.

Wasting and stunting often co-exist in children in LMICs and both are associated with increased mortality in childhood due to infectious diseases, particularly diarrhoea and pneumonia^35^. This susceptibility to infections is due to impaired immune function (including weakened gut-barrier function, humoral and cell mediated immunity) with consequent inadequate nutrient intake due to anorexia and malabsorption^36^. This further exacerbates immune suppression and impaired growth whilst energy and micronutrients are diverted to acute phase immune responses to combat multiple and often recurrent infections, leading to a chronic systemic inflammatory state and bacterial translocation^37^. Indeed, PEM is the primary cause of immune deficiency worldwide, and the vicious cycle of infection (clinical and sub-clinical) and PEM is well-described^38,39^.

In high income countries PEM is common among hospitalised adults, particularly the elderly, where 23-60% elderly patients in acute healthcare settings are malnourished^40^ and up to 50% of patients with concurrent morbidities are also affected^41^. The causes are commonly poor nutrient intake (for example, in the elderly due to poor oral health, depression, as a side effect of medication, or inadequate feeding support) and chronic underlying conditions that increase the metabolic demand due to inflammation, resulting in anorexia and increased muscle catabolism (cachexia), such as end stage renal failure^42,43^. This leads to altered body composition and adverse functional and clinical outcomes. The Global Leadership Initiative on Malnutrition has developed internationally validated diagnostic criteria based on both phenotypic (weight loss, low body mass index, reduced muscle mass/sarcopenia) and etiologic criteria (reduced food intake or assimilation, and inflammation or disease burden, including major infections or trauma) to facilitate early identification and management of patients with PEM to avert deaths and adverse outcomes^43^.

In the current SARS-CoV-2 global pandemic, there is an urgent need to identify PEM-related factors that render individuals vulnerable to succumbing to this infection. As a staggering 11% of the population are likely to have impaired immunity due to PEM^32^, many populations particularly in LMICs are potentially at risk of developing disease during this pandemic, although the severity of the trajectory is yet to be fully determined. Furthermore, although COVID-19 primarily affects the respiratory tract, patients can also have gastrointestinal symptoms including diarrhoea, nausea, and vomiting and loss of smell that can have an impact on nutrient intake and assimilation^44^. Human enteric coronavirus causes moderate to severe villous atrophy in animal models with virus particles visible in enterocytes of large and small intestine^45,46^. Coronavirus-like particles have also been found in degenerating jejunal epithelial cells of adults in India with histological evidence of malabsorption due to environmental enteric dysfunction and among Aboriginal children with lactose malabsorption post gastroenteritis^47,48^. However, the exact mechanisms of COVID-19 induced gastrointestinal symptoms of nausea, vomiting and loss of taste remain elusive^49^.

Although there is no current published data on the impact of PEM on the susceptibility and disease progression of SARS-CoV-2 infection in children, extrapolation from other RNA viral infections suggests that undernourished children are likely to have more severe respiratory and gastrointestinal disease. RNA viruses, including influenza A and B, and human metapneumovirus, are important pathogens causing pneumonia in children aged under 5 years globally^50^. PEM has been associated with influenza-related severe acute respiratory illness in under-5s in South Africa (adjusted odds ratio [aOR] 2.4; 95% CI, 1.1–5.6)^51^. In previous pandemics of influenza A (H1N1) such as the one in Guatemala in 2009 where 5 of the 11 deaths among hospitalised patients occurred in under 5’s, PEM was thought to have been a key contributing factor^52^. Children between 6 months and 5 years were thus identified as a priority group for vaccination^52^. However, to date children appear to be at lower risk of suffering severe episodes of COVID-19 than adults^53^.

### Systematic review

Our systematic search involved terms related to PEM in both children and adults and RNA viruses. The systematic screen of PubMed and EMBASE yielded 120 papers after removing duplicates; 23 were taken to full text screen and all were excluded as they did not examine the influence of PEM on coronavirus susceptibility or disease course.

A further search of the pre-print servers identified 3 studies that were included. Li *et al*. conducted a cross-sectional study and recruited 182 elderly hospitalised COVID-19 patients ≥65 years, in one centre in Wuhan, China^54^. The authors found that 53% were classified as malnourished using a mini nutrition assessment (based on recall of dietary intake) and 28% were at risk of malnutrition. There were no statistically significant differences in the triceps skin-fold thickness and mid-arm circumference between those who were non-malnourished, at risk of malnutrition or malnourished. However, diabetes mellitus (OR 2.12; 95% CI 1.92–3.21), low calf circumference (OR 2.42; 95% CI 2.29–3.53), and low albumin (OR 2.98; 95% CI 2.43–5.19) were independent risk factors for malnutrition. Their recommendation was for nutritional support to be enhanced for COVID-19 elderly patients with diabetes, low albumin and low calf circumference due to their increased risk of becoming malnourished. A retrospective study that included 141 COVID-19 patients in the analysis, explored the risk of adverse clinical outcomes among elderly patients (>65y) by nutritional status (using validated nutrition risk screening tools for adults including Nutrition Risk Screening 2002 (NRS-2002), Malnutrition Universal Screening Tool (MUST), Mini Nutrition Assessment Shortcut (MNA-sf), and Nutrition Risk Index (NRI)) in one hospital in China^55^. They found that patients at risk of PEM had significantly longer hospital stay, poor appetite, more severe COVID-19 disease and greater weight loss than patients not at nutritional risk using NRS 2002, MNA-sf, and NRI-2002. They recommended routine screening of elderly COVID-19 patients for nutrition risk coupled with nutrition interventions to improve clinical outcomes. Caccialanza *et al*.’s protocol is on a pragmatic trial in Italy for early nutritional supplementation with high-calorie dense diets combined with intravenous infusion of multivitamin, multimineral trace elements solutions for non-critically ill patients hospitalized for COVID-19 disease^56^. This was based on their observations of the drastic reduction in food intake due to severe inflammation among these patients at admission that predisposes them to poor respiratory outcomes. The nutritional interventions will be modified based on the clinical and nutritional status of patients during admission to include parenteral nutrition. The analysis of the effectiveness of this package of nutrition interventions is likely to be complex but is keenly awaited.

### Clinical trials

In the current pandemic, a similar pattern is being played out to what we have seen in previous pandemics. Patients with PEM, especially amongst the elderly and those presenting comorbidities, have been among those with the highest mortality^57^. Indeed a cross-sectional study in Wuhan, China with 182 COVID-19 patients aged ≥65y in a single centre found that diabetes (OR 2.12; 95% CI 1.92– 3.21), low calf circumference (OR 2.42; 95% CI 2.29–3.53), and low albumin (OR 2.98; 95% CI 2.43– 5.19) were independent risk factors for PEM^54^. Prolonged ICU admission causes or worsens existing PEM with associated sarcopenia (loss of skeletal muscle mass and function), exacerbated by the inflammation associated with the infection^58^. Identification and management of PEM is now a key component of managing patients with COVID-19 in Europe to avert adverse outcomes. There are no clinical trial data to guide the design of optimal nutrition management strategies in the context of COVID-19. The European Society for Clinical Nutrition and Metabolism has published nutrition rehabilitation guidelines primarily based on consensus and expert opinion using a combination of enteral and parenteral nutrition if oral intake not adequate^58^ (see also Section 16).

The clinical trials registry search identified 3 on-going studies in the US, Spain and France related to PEM, none of which are in children (Supplementary Material 4). All of these are observational studies. The US study (NCT04350073) seeks to undertake a detailed evaluation of the longitudinal energy expenditure and metabolic effects in COVID-19 adult patients, admitted to a single intensive care unit (ICU) with respiratory failure, using indirect calorimetry, cardiac assessment, body composition, and muscle and ultrasound measures. This is to guide the metabolic and nutritional care of these high-risk patients, optimise their care and ultimately improve outcomes. In Spain, the study (NCT04346212) seeks to assess the prevalence of oropharyngeal dysphagia among COVID-19 adult patients post discharge from one ICU and to describe their associated nutritional status, requirements for nutritional supplements and adaptations, in order to design strategies to optimise their care and clinical outcomes. The study in France (NCT04386460) seeks to explore the associated risks of dental health/isolation/ anorexia with malnutrition among elderly patients and evaluate the impact of dentists referring these at-risk patients to physicians on malnutrition prevention. The results of these studies are eagerly awaited as they will be key to informing the design of targeted nutritional interventions to both prevent and manage PEM in the context of COVID-19.

## 5. Overweight, obesity and diabetes mellitus

### Landscape review

Obesity is a recognised risk factor for type 2 diabetes mellitus (DM), and both have been associated with an increased burden of respiratory tract infections (RTIs)^59^. A systematic analysis found a U-shaped relationship between body size and risk of RTIs^60^ and DM has also been found to increase susceptibility to, as well as severity of, respiratory infections in general^61^. It is therefore not understood if they independently contribute to this increased morbidity and mortality risk^62^.

Obesity is causally related to, and potentiates, cardiovascular and metabolic derangements such as hyperglycaemia and DM^63^. This reduces the protective cardiorespiratory reserve and potentiates the immune dysregulation that appears, at least in part, to mediate the progression to critical illness and organ failure in a proportion of patients with severe respiratory infections including COVID-19^63,64^.

Several cellular mechanisms that may increase the susceptibility of DM patients to respiratory infections have also been described, including greater affinity of SARS-COV-2 for cell binding and entry, reduced viral clearance^65^, inhibited lymphocyte proliferative response to different kinds of stimuli^66^, as well as impaired monocyte/macrophage and neutrophil functions^67^.

### Systematic review

The systematic literature search yielded a total of 1331 articles; 947 were taken to title and abstract screen after 384 duplicates were removed. 115 articles were considered for full-text screening and 6 papers met the inclusion criteria for obesity and 12 for diabetes. The pre-print server search for obesity and diabetes yielded a total of 154 articles. 34 were considered for full-text screening and 29 of these met the inclusion criteria. Since included studies were numerous, and largely confirmed the same key messages of increased risk of severe disease progression, we did not extract all studies to Supplementary Material 5 but do refer to all included studies in the following narrative synthesis.

#### Obesity

Obesity is a frequent finding in hospitalised COVID-19 patients with the prevalence varying between studies: 10% in China^68^, 41.7%^69^ and 47.5%^70^ in the US and 75.8% in France^71^. A study compared 44 ICU COVID-19 patients in France with a historical control group of 39 consecutive acute respiratory distress syndrome patients admitted to the ICU just before the COVID-19 crisis and found obesity to be the most frequent comorbidity among patients (n=32, 73% vs n=11, 28% in controls; p < 0.001)^72^.

Obesity is generally associated with poor COVID-19 outcomes and this has been confirmed in all studies included in this systematic review. The contributory mechanisms, as has been suggested by Zhang *et al*.^73^, are aggravated inflammatory response, enhanced cardiac injury and increased coagulation activity. Their study which included 13 young patients who died of COVID-19 and 40 matched survivors found a higher body mass index among deceased individuals (OR = 1.35; 95% CI= 1.08-1.70)^73^. Another study has suggested that increased ACE2 expression in the bronchial epithelium of obese individuals may contribute to poor outcome^74^.

Obesity has been associated with higher risk of severe COVID-19 disease in many populations and across age brackets. A study by Cai *et al*. found that patients with a BMI >28kg/m^2^ had significantly higher odds of developing severe disease (aOR 3.40, 95% CI 1.40-2.86)^68^. Klang *et al*., in a study of 3406 patients, found poor outcomes in different age groups (young: <50y and old: ≥50y). For the younger population, BMI above 40 kg/m^2^ was independently associated with mortality (aOR 5.1, 95% CI: 2.3–11.1). For the older population, BMI above 40 kg/m^2^ was also independently associated with mortality but to a lesser extent (aOR 1.6, 95% CI 1.2 – 2.3)^75^. In a cohort of 46 pregnant women, 15 had severe COVID-19 with the majority being either overweight or obese (80%)^76^. Another study also found that obesity (BMI >30 kg/m^2^) was associated with increased risk of ICU admission or death (RR = 1.58, p = 0.002) whereas being underweight was not (RR = 1.04, p = 0.892)^77^.

Obese patients were more likely to require invasive mechanical ventilation, with severe obesity (BMI ≥35 kg/m^2^) found to be associated with ICU admission (aOR 5.39; 95% CI:1.13-25.64)^70^. Similar findings of adverse outcomes were found in other studies^71,78^. Hur *et al*. found that obese patients with COVID-19 had a decreased chance of extubation compared with non-obese patients^79^ [Hazard Ratio for extubation: 0.53 (95% CI: 0.32-0.90) for patients with a BMI of 30 to 39.99 and 0.40 (95% CI, 0.19-0.82) for those with a BMI of ≥40]. Palaiodimos *et al*. also found that severe obesity *i*.*e*. BMI ≥ 35 kg/m^2^ compared with those with a BMI 25-34 kg/m^2^, was independently associated with higher in-hospital mortality [3.78 (1.45–9.83)] as well as a significant predictor for intubation [3.87 (1.47– 10.18)]^80^.

#### Diabetes mellitus

Diabetes is a common comorbidity among COVID-19 patients and has been associated with poor outcomes in all included studies with the exception of Cariou *et al*. (see below). The frequency of diabetes among hospitalised patients was investigated in many studies, ranging from 3.8% in Iran^81^, 5.5%-35.7% in various studies from China^2,82–90^, 19.9% in the UK biobank^91^, and 33.8% in the USA^69^.

Hyperglycaemia in those with and without a history of diabetes may indicate a poor prognosis in COVID-19^92^. A study by Guo *et al*. suggests diabetes should be considered as a risk factor for a rapid progression and poor prognosis of COVID-19^93^. The utility of diabetes screening after admission has been suggested by Wang *et al*. who found high HbA1c level at admission to be associated with inflammation, hypercoagulability, and low SaO_2_ in COVID-19 patients^94^. This severe inflammatory response was also reported by other studies^93,95^. The mechanism, though not completely understood, may be through metabolic derangement such as that leading to ketosis and ketoacidosis. A study found that ketosis and ketoacidosis disproportionately affected diabetic patients compared with those without diabetes^90^.

Patients with diabetes are found to be more likely to develop severe or critical disease conditions with more complications, and had higher incidence rates of antibiotic therapy, non-invasive and invasive mechanical ventilation, and death (11.1% vs. 4.1%)^96^. Chen *et al*. found that diabetes and other factors such as increasing age, male sex and hypertension delay viral clearance thereby leading to a poor prognosis^97^. These risk factors are similar to those found in other studies^98–100^. COVID-19 patients with diabetes were more likely to develop severe or critical disease with more complications at presentation, and had higher incidence rates of antibiotic therapy, non-invasive and invasive mechanical ventilation and death (11.1% vs. 4.1%)^101^. In another study by Wu *et al*., the prevalence of diabetes among those with COVID-19-related acute respiratory distress syndrome (ARDS) was significantly higher than in those without ARDS (difference, 13.9%; 95% CI, 3.6%-24.2%)^102^. Bode *et al*. found in patients with diabetes and/or hyperglycaemia compared with those without, a longer median length of stay in hospital (5.7 vs 4.3 days, P <0.001) and higher mortality rate (28.8% vs 6.2%, p < 0.001)^103^. This mortality rate was similar to that found in another study (27.7%)^94^. Shi *et al*. found a higher proportion of intensive care unit admission (17.6% vs. 7.8%, p <0.01) and more fatal cases (20.3% vs. 10.5%, p <0.017) were identified in COVID-19 patients with diabetes than in the matched patients^104^. A study by Chang *et al*. found that patients with diabetes were more likely to progress to severe disease compared to those without [OR: 64.1 (4.6–895.5)]^105^. The findings were similar to those of Huang *et al* (OR: 4.3; 95% CI, 1.1-17.7)^106^. In Iran, Rastad *et al*. found diabetes alone or in association with other comorbidities was associated with increased risk of death [OR 1.69 (1.05–2.74) and 1.62 (95% CI 1.14–2.30) respectively]^107^. In a cohort of 28 diabetic patients, half required ICU admission^108^.

A study by Li *et al*. suggests that COVID-19 patients with newly diagnosed diabetes have a higher mortality risk of all-cause mortality [multivariable-adjusted HR: 9.42 (95% CI 2.18-40.7)] but this was not statistically significant compared with patients with normal glucose (1.00), hyperglycaemia [3.29 (95% CI: 0.65-16.6)] and known diabetes [4.63 (95% CI 1.02-21.0)]^109^. Increased mortality for patients with diabetes and COVID-19 has been linked to older age (aOR 1.09 [95% CI 1.04, 1.15] per year increase), elevated C-reactive protein (aOR 1.12 [95% CI 1.00, 1.24]) and insulin usage (aOR 3.58 [95% CI 1.37, 9.35])^110^. The latter finding on insulin use is in contrast to findings by another study which showed that patients with hyperglycaemia already treated with insulin infusion at admission had a lower risk of severe disease than patients without insulin infusion^111^. Metformin use, however, was associated with better outcomes in diabetics compared with those not receiving it^112^. These findings were complemented by Zhu *et al*. who found that well-controlled blood glucose (glycaemic variability within 3.9 to 10.0 mmol/L) was associated with markedly lower mortality compared to individuals with poorly-controlled blood glucose (upper limit of glycaemic variability exceeding 10.0 mmol/L) (adjusted HR: 0.14) during hospitalization^113^.

Only one study did not find diabetes to be associated with poor COVID-19 outcomes. Cariou *et al*. found that diabetes, HbA1c, diabetic complications and glucose-lowering therapies were not associated with disease severity (tracheal intubation for mechanical ventilation and/or death) within 7 days of admission^114^.

### Clinical trials

Searches of clinical trials databases revealed 13 planned or ongoing studies related to overweight/obesity or diabetes and COVID-19 (Supplementary Material 4). Of these nine were observational studies and four RCTs (two in the USA, one in Israel and one in Italy). Three of the RCTs evaluate the efficacy of the use of dipeptidyl peptidase 4 (DPP4) inhibitors (oral hypoglycemic agents: Linagliptin and Sitagliptin respectively) whilst another uses an antiviral nucleotide analogue (AT-527) on COVID-19 outcomes. All three studies using oral hypoglycemic agents evaluate their efficacy, compared with standard care, on clinical outcomes defined as lung disease in two studies and changes in glucose levels in one. The study using AT-527 seeks to assess its effect on progression to respiratory insufficiency, compared with a matching placebo, in moderate COVID-19 patients aged 45 to 80 years who are obese, or with a history of diabetes and hypertension.

## 6. Anaemia

### Landscape review

Anaemia is a condition where an individual’s haemoglobin concentration falls below the accepted lower threshold specific for their age, sex and pregnancy status. Anaemia remains highly prevalent worldwide, especially in low income countries, and particularly in South Asia and sub-Saharan Africa. The most common cause of anaemia worldwide is iron deficiency, which is caused by inadequate nutritional iron intake, impaired iron absorption, increased iron utilisation (for example during pregnancy or during rapid child growth), and blood losses (for example, menstrual blood losses, gastrointestinal bleeding, and blood donation). Anaemia is thus most common in preschool children, women of reproductive age, and during pregnancy^115^.

Beyond iron deficiency, there are many other causes of anaemia. During inflammation, iron may be withheld from the plasma through elevated hepcidin concentrations (functional iron deficiency); coupled with impairments on erythropoiesis and reduced red cell survival, this can result in anaemia of inflammation, which is common in patients with medical illnesses (such as cancer, infection and autoimmune conditions)^116^. Functional iron deficiency may also be an important component of the overall burden of anaemia in low income countries where exposure to endemic infections is intense. Other acquired causes of anaemia include haemolytic anaemias. These include autoimmune haemolytic anaemias, caused by autoimmune destruction of red blood cells (usually provoked by viral infections, some bacterial infections, underlying lymphoproliferative disorders, and medications)^117^. Other causes of haemolytic anaemia include microangiopathic haemolysis (which can be due to many causes including congenital, caused by infections, autoimmune conditions, cancer, pregnancy complications, and mediations). Bone marrow failure (aplastic anaemia, or replacement of the bone marrow by malignancy) can also cause anaemia. In the tropics a major cause of childhood anaemia is malaria, malaria anaemia has elements of haemolysis, marrow failure and functional iron deficiency. Other important causes of anaemia include genetic disorders of haemoglobin including alpha thalassaemia, beta thalassaemia and sickle cell disease.

Like all infections, acute viral infection can promote an innate immune response, elevation in hepcidin, and hence functional iron deficiency and anaemia of inflammation. Viral infections can also cause bone marrow failure. For example, Parvovirus B19 infection is frequently asymptomatic, or may cause a mild febrile illness with a rash (‘slapped cheek disease’). However, in immunocompromised individuals, and in individuals with chronic erythroid overactivity (e.g. haemolytic disease, sickle cell disease) it can cause cessation of erythropoiesis resulting in a transient aplastic crisis with severe anaemia. Parvovirus B19 during pregnancy can infect the fetus, causing failure of fetal erythropoiesis and severe fetal anaemia, which can result in hydrops fetalis and fetal death^118^.

### Systematic Review

From the PubMed and EMBASE database searches, after deduplication 407 articles were assessed at the title/abstract stage. Of those that mentioned anaemia we only considered those addressing potential nutritional causes of anaemia for formal data extraction, due to the scope of this review. However, several other types of anaemia featured in the initial screen, which we briefly summarise here. For example, two articles described the management of pernicious anaemia in the case of disrupted B12 treatment^119,120^. Two case series have provided preliminary information on beta thalassaemia major. A small series of 11 patients with beta thalassaemia in Italy infected with COVID-19 all experienced mild to moderate disease and all survived, even despite the presence of comorbidities associated with iron overload^121^. A nationwide study in Iran identified a lower incidence of diagnosed COVID-19 among patients with thalassaemia compared with the general population (8.7 per 10000 in the thalassaemia population compared with 11.0 per 10000 in the general population), although patients with thalassaemia may have been sheltering. Patients with thalassaemia experienced a higher mortality rate (26.6%) compared with the general population (6.3%); patients who did not survive had higher risks of comorbidities including diabetes, hypertension, and heart disease, although splenectomy was not a risk factor for mortality in this group^122^. A case report identified combined autoimmune anaemia (destruction of red blood cells by autoantibodies) and thrombocytopenia (destruction of platelets by autoantibodies) (collectively termed “Evan’s syndrome”) in a patient with COVID-19^123^. A case series from Belgian and French hospitals identified the onset of acquired warm and cold autoimmune haemolytic anaemia associated with a positive direct antiglobulin test in seven patients; four of the patients had a previous or new diagnosis of an indolent B cell malignancy, and viral infection may have triggered the onset of haemolysis^124^. These cases were each successfully treated using therapies including intravenous immunoglobulin, steroid and even rituximab, and all patients across these case series survived. There have been further case reports describing the association between autoimmune haemolytic anaemia and COVID-19^125,126^.

Whilst haemoglobin measurement has not been included in the core-outcome dataset proposed by WHO^127^, several studies suggest anaemia may be a clinical feature of COVID-19. For example, initial reports from Wuhan describing clinical features of COVID-19 pneumonia identified anaemia in up to 50% of patients whom mostly appeared to have severe disease^44^. A subsequent report from Wuhan identified anaemia in 15% of patients with COVID-19, with anaemia more common among non-survivors^2^. Similar haemoglobin concentrations have been reported in other COVID-19 cohorts^128^ and several studies include anaemia as a covariate in descriptive statistics. As in other medical conditions, anaemia appears to be associated with poorer prognosis, perhaps as a biomarker for more severe inflammation^129,130^.

After the title and abstract review nine articles were taken to full screen. Six articles did not address nutritional causes of anaemia. One paper by Cavezzi *et al*. was a review on the possible pathophysiological pathways by which SARS-CoV-2 may cause both haemoglobin dysfunction and hypoxia (through haemolysis and forming complexes with haem) and tissue iron overload (through mimicking the action of hepcidin)^131^.

Ultimately, we found two eligible studies for formal inclusion (Supplementary Material 5). The first was a case report of a patient testing positive for COVID-19 alongside several co-morbidities including severe iron-deficiency anaemia^132^. He was successfully treated with antiviral treatment alongside recombinant human Erythropoietin (rhEPO), leading the authors to propose further testing of the effectiveness of rhEPO in anaemic COVID-19 patients. The second study was a retrospective analysis of 259 patients hospitalised with COVID-19 in Austria^133^. The authors distinguished between those patients presenting with anaemia of inflammation at admission and those with iron-deficiency anaemia (IDA). Compared to patients with no iron deficiency, having IDA was associated with a longer hospital stay, but was not associated with increased mortality, risk of ICU admission, nor of mechanical ventilator use. However, when considering purely anaemic versus non-anaemic patients, the anaemic patients had a higher risk of death (OR 3.729 (95%CI 1.739– 7.995). Of these anaemic patients, the majority (68.8%) had anaemia of inflammation, which the authors describe could be linked to co-morbidities, or to the advanced inflammation associated with COVID-19, or both. Collectively, these limited data indicate anaemia is an adverse prognostic indicator in severe COVID-19.

From the pre-print server screen, of the 122 articles returned 4 were taken to full screen review and none were eligible.

### Clinical trials

The search of clinical trial registries did not identify any ongoing clinical studies specifically evaluating the effects of anaemia, or treatment of anaemia, on COVID-19 prognosis.

## 7. Iron

### Landscape Review

Approximately 2% of human genes encode proteins that interact with iron, and around 6.5% of enzymes depend on iron^134^. Viruses co-opt host cellular processes to replicate, so it is unsurprising that viral replication utilizes proteins that are iron-dependent^135^, such as ribonucleotide reductase (the key enzyme involved in nucleotide biosynthesis). Consequently, viral pathogenesis could be influenced by cellular iron status. However, several features of host responses to viral infection could also be affected by iron, for example macrophage polarisation and lymphocyte proliferation, potentially influencing either disease susceptibility or course.

Iron deficiency is the most prevalent micronutrient deficiency worldwide, most prominently causing anaemia. The major burden of iron deficiency is borne by young children and women of reproductive age - groups at lower risk of COVID-19 mortality^136^ - and pregnant women (for whom patterns of COVID-19 hospitalisation risk appear similar to the general population^137^)^138^. Functional iron deficiency, where iron is present but sequestered and unavailable in circulation, occurs during many chronic conditions, including obesity^139^ – a known COVID-19 risk factor^136^.

Effects of iron status on infection susceptibility are not fully defined, and likely vary according to age, setting (e.g. malarial or non-malarial) and type of infection^140,141^, meaning caution should be employed in making extrapolations to viral infections in general and specifically to COVID-19. Iron deficiency protects against certain microbial infections including malaria^142^, and iron supplementation exacerbates malaria risk in children in malaria-endemic areas in the absence of malaria control measures^143,144^. Excess iron increases siderophilic bacterial infection risk^145^, and elevated iron indices predict mortality during HIV-1 infection, even after adjustment for CD4 count and inflammation^146^. Non-malarial infections, including gastrointestinal and respiratory infections, are also reported in several trials of childhood iron supplementation^144^. One large intervention trial in Pakistan reported increased signs of respiratory infection in children administered iron^147^, although other smaller trials have reported contrasting effects of iron supplementation on incidence of respiratory tract infections in children^140,148–150^. However, high quality evidence on interactions between iron status or interventions and specific respiratory viral infections in humans is lacking.

Although precedents from other human viral infections are limited, iron could in principle affect several aspects of the host-SARS-CoV2 interaction:

- As discussed above, viral replication, in general terms, co-opts several iron-dependent host cellular processes^135^.
- Impaired lung function and hypoxia are key features of severe COVID-19 disease, and iron deficiency exaggerates the pulmonary response to hypoxic stress^151,152^.
- Iron levels may influence macrophage polarisation and cytokine production^153^, potentially influencing COVID-19-related inflammatory phenotypes.

In addition, a rare mutation of *Tfrc* (encoding the transferrin receptor) that disables cellular iron uptake causes severe combined immunodeficiency in children^154^. Nutritional iron deficiency or pre-existing functional iron deficiency have also been linked to immune impairment^155^. Moreover, during many infections, interleukin-6 (IL-6)-mediated stimulation of the iron regulatory hormone hepcidin, as part of the hepatic acute phase response, causes macrophage iron sequestration and acute reduction in serum iron concentration^141^. Common respiratory infections and fevers associate with hepcidin upregulation in African children^156^. A key feature of COVID-19 severe/critical disease is excessive production of inflammatory cytokines, notably IL-6, and accordingly, raised hepcidin has been reported in hospitalised COVID-19 patients^157,158^. Consistent with involvement of hepcidin activity, extreme hypoferraemia has been reported in several studies in severe COVID-19 patients, with serum iron concentration shown to be highly predictive of disease severity^157,159–161^. A further retrospective analysis (also described in Section 6 on anaemia) also reported perturbed markers of iron homeostasis in hospitalised COVID-19 patients, with functional iron deficiency classified in approximately 80% of patients at admission^133^. Whether or not this functional iron deficiency limits the development of the adaptive response (analogous to the effect of the *Tfrc* mutation^154^) in the context of SARS-CoV-2 infection remains to be determined.

### Systematic review

Besides “iron”, our systematic search involved terms, related to common biomarkers of iron status and iron handling – including “ferritin”, “transferrin”, “Tsat” [transferrin saturation] and “hepcidin” (Supplementary Material 2). The systematic screen of PubMed and EMBASE returned 110 papers after removing duplicates; 45 were taken to full text screen, all of which were excluded as none examined the influence of iron deficiency or interventions on coronavirus susceptibility or disease course.

A further 10 distinct studies were identified through the pre-print server screen; again, all were excluded for the same reasons. The combined screen of PubMed/EMBASE and pre-print servers did identify 32 original studies or meta-analyses reporting effects of coronavirus infection on iron-related markers, most prominently the iron storage protein ferritin. However, in the context of typically extreme COVID-19 associated inflammation, serum ferritin is not useful as a marker of iron status, yet it does show relevance as an indicator of disease severity and could potentially reflect iron dysregulation besides inflammation (see Box 2).

### Clinical trials

The clinical trial screen returned 134 trials. Amongst these 124 were identified owing to inclusion of ferritin concentration amongst clinical outcomes. No clinical trials pertaining to iron supplementation, or investigations of baseline iron status on COVID-19 susceptibility or progression were identified. However, three clinical trials were identified aimed at targeting iron during COVID-19 infection (Supplementary Material 4): each proposes to examine the effect of deferoxamine (Desferal^®^) on COVID-19 disease course and mortality, an approach discussed in a recent review^162^. The rationale was not described in two of the three trials; in the third, a rationale of reducing iron-induced lung toxicity was proposed. Iron chelation can reduce replication of viruses including HIV-1 in vitro^162^, yet its effect on viral pathogenesis in vivo is less clear. Given the emerging importance of iron in immune function and the uncharacterised role of iron in the SARS-CoV2 life cycle, outcomes of trials of iron sequestration in the context of COVID-19 are awaited with interest.

#### Box 2

**Ferritin and COVID-19**

Ferritin was included as a systematic review search term since low serum ferritin is frequently used in diagnosis of iron deficiency^163,164^. The initial screen returned 22 papers, 9 preprints and 124 clinical trials mentioning ferritin, none of which related to iron status assessment. Instead, elevated ferritin (hyperferritinaemia) was consistently reported in COVID-19 patients, with levels highest in critical disease (see meta-analyses^165,166^). While serum ferritin is upregulated in response to increased iron, it is also induced during inflammation by IL-1β and TNF-α, often correlating with inflammatory markers such as C reactive protein (CRP); as such, inflammation is a well-known confounder of ferritin-based iron status assessment^163,167^. Given that severe COVID-19 disease is characterised by hyperinflammation, reminiscent of other syndromes with macrophage activation-related hyperferritineamia^167,168^, serum ferritin levels will not reflect iron levels in the majority of COVID-19 patients. However, it does show potential as a prognostic biomarker given its association with COVID-19 disease severity^165,166^. Whether or not ferritin plays an active role in disease pathogenesis, or merely reflects the degree of inflammation and macrophage activation warrants further attention.

## 8. Vitamin A

### Landscape review

Vitamin A has an established role in supporting immune function and protecting against viral infections. Evidence from animal studies shows clear effects of serum retinol level on mucosal immune function and intestinal lymphocyte action, and protection against viral infections of the respiratory and intestinal tracts^169–173^.

The effectiveness of viral vaccines is compromised by low serum vitamin A through the suppression of immunoglobulin G1 (IgG1)^172,174^ and inflammatory responses^173^. Vitamin A also modulates other immune components through its action on dendritic and natural killer cells^175^. It is essential in maintaining epithelial tissue integrity^176^, which is severely damaged in viral infections such as measles^177^. Recent systematic reviews conclude that vitamin A supplementation in children is associated with a reduction in all-cause mortality, and with reductions in the incidence of measles and diarrhoea, but there is little evidence to support a beneficial effect on respiratory infections^178,179^.

Serious COVID-19 caused by SARS-CoV-2 infection has some similar manifestations to measles including fever, cough and pneumonia (though it is important to note that the severe lung pathology of COVID-19 has a distinct pathophysiology from other viral pneumonias)^180^. People with underlying chronic diseases and impaired immunity are also at high risk for both COVID-19^181,182^ and measles^183^.

Vitamin A is recommended by the World Health Organization as part of the standard treatment package for all children with acute measles^184^. The COVID-19 pandemic has likely increased measles mortality – more than 20 countries have suspended measles vaccination and vitamin A supplementation campaigns as healthcare workers focus attention on COVID-19 leading to a surge in measles infections and mortality particularly in low income settings such as the DR Congo where measles has killed more than 6500 children and is still spreading^185^. Vitamin A is recommended mainly to reduce mortality^186^ and risk of complications from pneumonia, croup and ocular problems^187^ by correcting the low or depleted retinol levels resulting from measles infection. The treatment regimen consists of the administration of high dose vitamin A on two consecutive days. Children with evidence of deficiency (ocular symptoms) receive a repeated dose at 2 to 4 weeks^184^. A Cochrane systematic review of eight trials^188^ and another systematic review of six trials^189^ showed no overall reduction in mortality with vitamin A treatment of measles. However, when stratified by vitamin A treatment dose, administering two doses (on consecutive days) reduced measles mortality significantly in both meta-analyses with RR=0.38 (95% CI 0.18-0.81)^188^ and RR=0.21 (95% CI 0.07-0.66)^189^, and therefore forms the basis for the recommended regimen of vitamin A treatment of measles.

A recent non-randomised study observed a reduction in mortality among 330 Ebola virus patients who received vitamin A supplementation compared to 94 patients who, due to supply problems, did not receive vitamin A (RR=0.77 (95% CI 0.59-0.99))^190^. This trial is limited by significant risk of confounding.

### Systematic Review

The systematic search of PubMed and EMBASE databases yielded 44 articles. After removal of duplicates (n=5) and those not meeting inclusion criteria (n=36), 3 systematic review articles were considered for full text extraction to examine reference lists for potentially eligible articles. No papers were included from examining reference lists. Our preprint search on vitamin A and COVID-19 yielded one potential paper which did not meet the inclusion criteria.

### Clinical trials

Two small sized randomized clinical trials involving vitamin A in the treatment of COVID-19 patients were identified from the clinical trials registries search. One of the trials, targeting 30 hospitalised patients (15 in the intervention arm) involves the use of an oral nutrient supplement (anti-inflammatory/antioxidant nutrients and vitamins) as supportive care for COVID-19 and includes 2840 IU vitamin A among other nutrients in the supplement for 14 days (NCT04323228). The reason for using anti-inflammatory or anti-oxidant nutrients in COVID-19 patients in this trial is to modulate the cytokine storm associated with the disease on the lungs. The other trial, targeting 80 hospitalised (non-ICU) patients (NCT04360980) uses an unspecified amount of vitamin A as part of a combination of nutrients given to the control group or standard of care (n=40).

## 9. Vitamin C

### Landscape Review

Vitamin C (ascorbic acid), synthesised by all mammals except humans and guinea pigs, supports diverse aspects of immune function by strengthening epithelial barriers, enhancing the function of adaptive and innate immune cells, promoting cell migration to infection sites, and participating in macrophage microbial killing^191^.

Unfortunately, vitamin C has a particularly chequered history in relation to viral infections. Double Nobel Laureate Linus Pauling blighted the end of his career by promoting mega-doses of vitamin C as a cure for common colds^192^ and cancers^193^ despite an absence of any robust evidence. Even today it is difficult to interpret the scientific and allied literature without encountering partisan opinions, and there remains a widespread popular view that vitamin C is effective. Pauling’s favoured mechanism of action was through its anti-oxidant effects. His belief in, and self-medication with, mega-doses of vitamin C runs contrary to the fact that there is a renal threshold leading to diminished retention and tissue saturation at oral intakes above 200mg/d^194,195^. Intravenous infusion of large doses of vitamin C can elevate leukocyte levels much further, but the putative mechanism of action against cancers (as yet unproven in humans) is proposed to be through its pro-oxidant effects of generating hydrogen peroxide at large doses^196^. This is pertinent to the on-going therapeutic trials in COVID-19 patients listed below.

Regarding the common cold, the most recent Cochrane review^197^ summarised 24 trials with 10,708 participants and found no evidence in the general population that regular consumption of vitamin C at 200mg/d or above reduced the incidence of colds (RR = 0.97 (95%CI 0.94 – 1.00)). In contrast, five trials with 598 marathon runners, skiers and soldiers on subarctic exercises yielded a combined RR of 0.48 (95%CI 0.35 – 0.64). The possibility that free radicals generated by extreme exercise are quenched by vitamin C provides a plausible explanation for this heterogeneity of results. Thirty-one trials covering 9745 episodes showed that taking regular vitamin C shortened the duration of symptoms in adults by 8% (95%CI 3 – 12%) and in children by 14% (95%CI 7 – 21%). Seven trials of therapeutic use of vitamin C administered at the start of an infection in 3249 episodes revealed no evidence of altered duration or severity. A single additional RCT in 1444 Korean soldiers has been published since the meta-analysis and reported a marginally significant reduction in incidence of colds among soldiers receiving 6000mg/d vitamin C orally (RR 0.80, 95%CI 0.64 – 0.99)^198^.

A Cochrane meta-analysis of the potential effect of vitamin C on the prevention and treatment of pneumonia has been updated very recently^199^. The results from 7 studies (5 RCTs and 2 quasi-RCTs) involving 2774 participants (children, adults, army personnel) receiving doses ranging from 125 to 2000 mg/d vitamin C were judged to provide very low-quality evidence with respect to both prevention and treatment; hence no conclusions can be securely drawn.

For critically-ill patients the prior evidence for efficacy of low-to moderate-dose vitamin C (alone or as a cocktail with other anti-oxidants) is weak. A recent systematic review and meta-analysis of 11 RCTs found no evidence of benefit for mortality (9 trials) or any secondary outcomes^200^. There was a non-significant tendency towards mortality reduction in subgroup analysis confined to intravenous administration of high-dose vitamin C^200^. The meta-analysis was dominated by a large and robust multi-centre trial of 1223 ICU patients with half randomised to anti-oxidants including 1500mg/d enteral vitamin C (with or without glutamine) which reported no effect on survival (primary outcome) or on any secondary outcomes^201^.

The evidence from prior trials of high-dose intravenous vitamin C (HDIVC) in pneumonia and ARDS-type conditions is also of low quality and was either not summarised, summarised poorly, or in a biased manner in most trial registrations. One reason for the high interest in intravenous vitamin C can be traced to a single-centre uncontrolled observational study of 94 sepsis patients that reported a 5-fold reduction in mortality when vitamin C and thiamine were combined with hydrocortisone^202^. A follow-up multi-centre RCT of the same regimen in sepsis patients (the VITAMINS Study) has very recently reported no benefit in any outcome^203^. The CITRIS-ALI Trial in 7 US ICUs randomised 167 patients with sepsis or ARDS to 200mg/kg/d intravenous vitamin C or placebo for 4 days. There was no difference in the primary outcome of Sequential Organ Failure Assessment score or in the secondary outcomes of CRP or thrombomodulin^204^. In un-prespecified exploratory analysis not adjusted for multiple testing there was some evidence of enhanced survival to 28 days.

### Systematic review

From a total of 54 papers returned, 4 papers were identified for full screen. Most papers were commentaries or non-systematic reviews. In no case was there any substantive new data on clinical outcomes. Two papers used a systems biology bioinformatic approach to explore mechanisms through which vitamin C might be active^205,206^.

The search of preprint servers yielded 13 relevant papers all of which were accessed for full review; most were commentaries or editorials. Two systematic reviews concluded that the evidence that vitamin C was likely to benefit COVID patients was weak or absent^207,208^.

### Clinical trials

The search of clinical trials registers in June 2020 yielded 27 entries involving vitamin C. Three were observational studies, and 8 used vitamin C as a placebo (reportedly because vitamin C tablets are a similar size and appearance to the hydroxychloroquine tablets used in all these trials). Of the remaining 16 trials where vitamin C was, or was part of, the active compound under test, 2 did not clearly state dose or mode of administration. Four trials involved dietary supplements of vitamin C combined with other micronutrients, herbal remedies or in one case methylene blue and n-acetyl cysteine. These trials target 1220 participants at various stages of SARS-CoV-2 infection; usually in mild disease or testing the prophylactic value in healthcare workers. Based upon prior trials of HDIVC in patients with pneumonia, sepsis and cancers, 10 trials involve the intravenous administration of vitamin C.

The 10 currently-registered trials of HDIVC for COVID involve a target of 2,758 adult patients hospitalised with significant-to-critical COVID disease. They range from Phase 1 to 4. Three studies involve single-day bolus treatments with 10-20g vitamin C (for a 70kg individual). The remaining studies use doses ranging from 14 to 66g per day over 3-8 days with total doses amounting to between 56 and 327g of vitamin C (again for a 70kg individual). The rationale for these mega-doses is mixed, with claims of both anti-oxidant and pro-oxidant mechanisms, sometimes within the same rationale statements. Note that these doses are between 150 and 730 times higher than the recommended daily intake, and 7 to 33 times higher than the US Institute of Medicine’s Tolerable Upper Limits for vitamin C^209^. These should be viewed as pharmaceutical trials having no reference to vitamin C’s normal physiological functions. Based upon the paucity of prior evidence the investment in such trials is questionable.

## 10. Vitamin D

### Landscape Review

The wide-spread distribution of the vitamin D receptor (VDR) and vitamin D-metabolising enzymes in cells and tissues, including those of the immune system, is evidence of a wide-role for vitamin D in health. The role of vitamin D in the immune system has been reviewed recently^210,211^, including in relation to COVID-19^212–215^, and spans aspects of the immune system including the maintenance of barrier defences, innate immune response and an immunoregulatory role in antigen presentation and the adaptive immune responses^210,216,217^. As part of the innate immune response, antimicrobial peptides play an important role in the first line of defence against infections, including in respiratory infections^218^. Vitamin D is required for the production of anti-microbial peptides such as cathelicidins in macrophages and in the epithelial cells of the airways^217^ and in an RCT, vitamin D supplementation was shown to increase levels of antimicrobial activity in airway surface liquid^219^. Vitamin D can also reduce the production of pro-inflammatory Th1-type cytokines^210,212^ that are implicated in the cytokine storm associated with more serious COVID-19 clinical outcomes such as acute respiratory distress syndrome and multiple-organ failure^212,220,221^. The binding site for SARS-CoV-2 is ACE2^222^. Studies have shown that higher levels of ACE2 can reduce acute lung injury from infection and that vitamin D can modulate the expression of enzymes balancing the expression of ACE2 and ACE (reviewed in^223–225^) providing a mechanism for a potential role for vitamin D in the prevention and progression of COVID-19. 25OHD concentration may decrease as part of the acute phase response so data from observational studies in acutely ill patients should be interpreted with a degree of caution^226–228^.

Vitamin D deficiency (VDD) is prevalent across all continents, not only those at more extreme latitudes^229–232^ and certain groups are at particular risk including the elderly (especially those in care homes), ethnic minorities (living at higher latitudes) and the obese. There is a strong overlap between groups at risk of COVID-19 morbidity and VDD (ethnic minorities, obese, institutionalised elderly). Groups identified at higher risk of serious illness with COVID-19^233^ are also at risk for VDD, not only from low circulating 25OHD per se, but also lower circulating vitamin D binding protein (DBP), e.g. in patients with renal or hepatic disease^234^.

Human data from both observational studies and intervention trials support a role for vitamin D in the prevention of respiratory infections. Meta-analyses of observational data have found associations between low vitamin D status and both risk of acute respiratory infection^235,236^ and severity of symptoms^236^. A meta-analysis^237,238^ of individual participant data found a reduced risk of acute respiratory infection (aOR (95% CI): 0.88 (0.81 – 0.96)), particularly in individuals receiving regular (weekly or daily) vitamin D supplementation and in those with baseline 25OHD < 25 nmol/L (0.30 (0.17 – 0.53). More recent trials of respiratory infection prevention in children and adults have reported both a beneficial^239–241^ and no effect^242–245^ of vitamin D supplementation. The findings from a recently published large trial (n 5110) in New Zealand found no effect of a bolus dose of vitamin D on the incidence of acute respiratory infection^246^. The results of another large trial in 25,871 men (≥50 y) and women (≥55 y) of vitamin D and/or omega-3 fatty acids found no reduction in all-cause mortality whilst results for respiratory conditions are yet to be published^247,248^

Genetic polymorphisms within the genes for DBP, vitamin D-metabolising enzymes and the VDR may affect vitamin D transport, metabolism and action. Polymorphisms within the DBP have a small effect on DBP and 25OHD concentration^249^ and metabolism^250^ as well as response to supplementation^251,252^. VDR polymorphisms may impact the risk and progression of disease although results are mixed^253,254^. A recent meta-analysis in relation to enveloped-virus infection (a group that includes coronaviruses) found significant associations between certain VDR polymorphisms and susceptibility to respiratory syncytial virus^255^.

### Systematic review

From a total of 59 papers returned from Pubmed and Embase searches, 9 were taken to full text screen and two papers^224,256^ were identified for full screen. D’Avolio *et al*. found that mean 25OHD concentration measured a median 3 days after a COVID-19 PCR test was lower in 27 PCR-positive patients compared with 80 PCR-negative patients (28 vs 62 nmol/L; P=0.004)^256^. In an ecological analysis, Ilie *et al*. observed an inverse correlation between both COVID-19 case numbers and mortality figures against published population mean 25OHD concentrations (both r = −4; p=0.05) across 20 European countries^224^.

Screening of pre-print servers revealed a total of 38 manuscripts after exclusion of those previously identified from the Pubmed/Embase search. Of these, six were taken to full review.

Manuscripts described observational studies and investigated 25OHD concentration in COVID-19 positive cases. Three studies had fewer than 20 participants with both COVID-19 and vitamin D test results, and no control group; 2 reports measured 25OHD concentration in hospital in-patients: Cunat *et al*. reported 13/17 intensive care unit patients had 25OHD concentration less than 31 nmol/L^257^ whilst Lau *et al*. found that 11/13 ICU patients had 25OHD < 75 nmol/L compared to 4/7 in-patients, although there was no significant difference in mean 25OHD concentration between groups^258^. A third report from Indonesia in 10 hospitalized COVID-19-positive patients, found that 9/10 had a 25OHD concentration less than 50 nmol/L and 4/10 less than 25 nmol/L^259^.

A larger Belgian study described lower 25OHD concentrations and greater prevalence of VDD (defined as < 50 nmol/L) in a group of hospitalized COVID-19 patients (n 186) compared with a group of 2717 patients of similar age distribution sampled a year earlier (47 nmol/L and 54 nmol/L, p=0.0016; 59% vs 45%, p=0.0005). However, when stratified by sex, the significant difference in 25OHD concentration and VDD only remained in males^260^. In a study of 499 hospitalised patients or health care workers in the USA (Chicago) with a COVID-19 test result and vitamin D status measurement (in the past year) there was no difference between COVID-19 positive and negative cases (p=0.11)^261^. An expanded analysis that sought to categorize the vitamin D status of an individual based on (1) their vitamin D status test result and (2) vitamin D treatment regimen in the previous 2 years found that participants who were predicted ‘vitamin D deficient’ had an increased risk (relative risk = 1.77, p<0.02) of testing positive for COVID-19 compared with participants with predicted vitamin D status of ‘likely sufficient’^261^. In a different approach, Haustie *et al*. used baseline UK Biobank data from 348,598 participants collected 10 to 14 years ago of whom 449 had a positive COVID-19 test in between March and April 2020. After inclusion of other factors such as season, ethnicity and other health conditions there was no significant association between 25OHD and COVID-19 infection (OR = 1.00; 95% CI = 0.998 - 1.01)^262^.

Two additional studies were identified from reference screening. A study from the Philippines found that in 212 COVID-19 hospitalized patients, vitamin D status was associated with clinical outcomes such that for each standard deviation increase in 25OHD concentration, the odds of having a mild clinical outcome rather than a severe or critical outcome were 7.94 and 19.61, respectively (CI not reported)^263^. A study of 780 COVID-19 positive hospital patients found that after correction for age, sex and comorbidity the odds ratio of death was 10.2 p<0.0001 (95% CI not reported) in cases with VDD (defined as < 50 nmol/L) compared with ‘normal’ vitamin D status (defined as 75 nmol/L)^264^. However, this study has since been discredited^265^.

### Clinical trials

Searches of clinical trials databases revealed 21 planned or ongoing studies related to vitamin D and COVID-19. Of these four were observational studies. The remaining 17 focussed on treatment (including disease progression) (n 12), prevention (n 2) or both prevention and treatment (n 2). Of the four prevention studies, vitamin D is registered as the main intervention for one trial, whilst two use vitamin D as an adjuvant with hydroxycholoroquine and one as a placebo. Of the remaining 13 trials, four use vitamin D in all groups, two as an adjuvant to the main treatment and seven either vitamin D3 (between 25 µg daily to single, bolus dose of 10 mg), vitamin D2 (1.25 mg twice weekly) or 25OHD (0.266 mg daily) as the primary intervention (one in combination with zinc). Study size ranges from 64 to 3140 participants.

## 11. Vitamin E

### Landscape review

Vitamin E is the collective term for 4 tocopherols and 4 tocotrienols^266^. Human dietary requirements are based on α-tocopherol, but there is increasing evidence of biological functions for the related compounds, including in relation to immunity^267^. Vegetable oils and nuts are rich sources of vitamin E and hence human deficiency is rare; thus the interest in vitamin E and immunity is frequently related to the question of whether supplementary vitamin E might improve immunity in at-risk subgroups such as smokers or the elderly.

The main biological role of vitamin E is as an anti-oxidant that quenches oxidative cascades especially of membrane poly-unsaturated fatty acids (PUFAs) in which it is highly soluble and hence penetrant^266^. Animal, human and cell culture studies have examined the role of supplemental vitamin E on a wide range of innate and adaptive immune cells. Numerous possible mechanisms of action are postulated (maintenance of cell membrane integrity, increased (and decreased) cell proliferation, increased IL-2 and decreased IL-6 production, enhanced immunoglobulin production, etc) but few confirmatory studies are available^266,267^.

Due to their dual and overlapping roles in antioxidant pathways there are close parallels between selenium and vitamin E with regard to immune function; roles that have been best studied in regard to viral infections. In the section on selenium, we describe the work by Beck and her team demonstrating that the virulence of coxsackie B3 and influenza H3N2 viruses is enhanced in selenium deficient hosts resulting from systematic viral mutations (see section 13). Beck’s team have used the same mouse protocol with vitamin E deficient mice and demonstrated that the viral mutation and enhanced pathogenicity is recapitulated with either or both selenium and vitamin E deficiency^268–271^, an effect that is enhanced in iron-loaded animals due to the increased oxidant stress.

The evidence for interactions between vitamin E status or supplementation and viral infections in humans is sparse and there are no available meta-analyses as a consequence. A recent (non-systematic) review has tabulated summary outputs from 8 studies of human infections of which 5 relate to respiratory infections^266^. Several of the studies involved post-hoc sub-group analysis of smokers and hence have questionable validity and poor generalisability^272,273^. The best study was a 2×2 factorial design of multivitamin-mineral or vitamin E supplementation in free-living adults >60 years old^274^. In 652 participants with 1024 respiratory infections there was no benefit of either regime in reducing incidence, and some evidence that vitamin E made the infections more serious^274^.

### Systematic review

Results from the systematic literature review for vitamin E are highlighted in Supplementary Material 3. From a total of 39 papers returned, 9 duplicates were removed and 30 titles and abstracts screened. Six review papers were considered for full text screen and to check reference lists for possible papers. None had substantive novel relevant information.

The search of preprint servers yielded four papers of which two were accessed for full review; these were both general reviews and lacked substantive new information in relation to coronaviruses or severe ARDS^207,208^.

### Clinical trials

The search of clinical trials registers yielded a single entry (NCT04323228) involving a very small study (n=30) in Saudi Arabia with vitamin E administered to 15 patients as part of a broad antioxidant cocktail.

## 12. Poly-unsaturated fatty acids (PUFAs)

### Landscape review

Long-chain poly-unsaturated fatty acids (LC PUFAs) are classified into two series (ω-3 or ω-6) according to the position of their double bonds. Both series have extensive immunomodulatory activity with ω −3 PUFAs tending to be anti-inflammatory and ω-6 PUFAs tending to be pro-inflammatory. ω −3 fatty acids are abundant in fish oils and ω-6 in vegetable oils. The ω-3 and ω-6 synthetic pathways compete for the same elongase, desaturase and ω-oxidation enzymes and hence the ratio of ω-3 to ω-6 series can be especially crucial. Comprehensive reviews of the immunomodulatory effects of PUFAs are available elsewhere^275–280^.

In brief, LC PUFAs exert immunomodulatory effects through a number of generic mechanisms. Eicosapentaenoic acid (EPA; ω-3) and arachidonic acid (ARA; ω-6) are precursors of eicosanoids; ARA generates inflammatory-type eicosanoids and EPA-derived eicosanoids tend to be anti-inflammatory^277,279^; a property that may be crucial to COVID-19 disease (see below)^276^. When incorporated into cell membranes LC PUFAs can beneficially modulate the activity of T-cells and other components of cellular immunity^279^. They also modulate cytokine responses; with ω-3 fatty acids tending to enhance IL-10 and suppress IL-6 production as well as inhibiting NF(κB)^279^. More recently PUFAs have been shown to play a crucial role in the production and action of specialised pro-resolution mediators (SPMs) that play a crucial role in ending the inflammatory cycle and thereby avoiding an excessive inflammatory response and cytokine storm. EPA and DHA (docosahexaenoic acid; ω-3) are precursors for resolvins and DHA is the precursor for protectins and maresins^276^.

Despite the wealth of biochemical evidence for key roles of ω-3 PUFAs in anti-inflammatory pathways the evidence of clear roles in human health is less robust. Meta-analyses with a range of health outcomes have failed to provide evidence for efficacy and in those where efficacy seems secure it is usually only achieved at high doses.

There have been several meta-analyses of the effects of ω-3 fatty acids from fish oils on critically ill patients. Due to differences in selection criteria and outcome measures the outcomes are varied. In 2018, Koekkoek *et al*.^281^ reviewed 24 RCTs of fish-oil containing enteral nutrition involving 3574 patients. There was no significant benefit on the primary outcome of 28d, ICU or hospital mortality. However, fish-oil administration significantly reduced length of stay (LOS) in ICU and duration of ventilation. In a pre-planned subgroup analysis there was a reduction in 28d mortality (OR 0.69, 95%CI 0.54-0.89), ICU LOS (−3.71 days, 95%CI −5.40 - −2.02) and duration of ventilation (−3.61 days, 95%CI −5.91 - −1.32) in patients with acute respiratory distress syndrome (ARDS). In 2019, Langlois *et al*.^282^ conducted a meta-analysis of the RCTs of ω-3 PUFA administration on gas exchange (PaO2-to-FiO2) and clinical outcomes in 12 trials involving 1280 ARDS patients. There was a significant early increase in PaO2-to-FiO2 that diminished but remained significant at days 4-7. There were non-significant trends towards reduced ICU LOS and duration of ventilation but not improvement in mortality, length of stay in hospital or infectious complications. Also in 2019, Dushianthan *et al*.^283^ meta-analysed 10 RCTs of enteral ω-3 supplementation in a total of 1015 ARDS patients. There was no benefit to all-cause mortality (OR 0.79, 95%CI 0.59 – 1.07) or any of the secondary outcomes. All of these meta-analyses encountered studies with high risk of bias and poor-quality evidence.

### Systematic review

From a total of 37 papers returned, 5 were taken to full screen, and none yielded relevant information not already considered.

The search of pre-print servers yielded one paper^284^ that extensively reviews the role of inflammation and the cytokine storm in lung damage but cites no supportive evidence for a modulating role of PUFAs other than that already reviewed above.

### Clinical trials

Notwithstanding this rather weak evidence of benefit in critically-ill patients including those requiring ventilation there have been calls for clinical trials of intravenous high-dose fish-oil lipid emulsions (FOLE) in hospitalized COVID-19 patients^285,286^. The first of these recommends use in patients at special risk of hyperinflammatory outcomes (e.g. the obese)^285^. In the second call, Torrinhas *et al*. emphasise the need to tailor dosage to body weight, recommend its use in all patients and that it should be combined with aspirin^286^. A very comprehensive summary of the putative benefits of high-dose fish oil has recently been published^276^. Despite these calls for intravenous FOLE trials, none have yet been registered.

The search of trial registers yielded three trials that had PUFA in the active intervention arm (see Supplementary Material 4). The two trials in hospitalized patients in the USA are testing eicosapentaenoic acid (EPA). One of the trials combines the EPA with docosahexaenoic acid (DHA) and gamma linolenic acid (GLA) plus additional antioxidant micronutrients. The third trial, in Latin American countries, will test whether icosapent ethyl (IPE) will prevent occurrence of COVID in at-risk health providers. Severity of disease will also be compared against placebo.

## 13. Selenium

### Landscape review

There is very strong evidence that selenium, through its role as a cofactor in the two key anti-oxidant pathways in humans (reduction of glutathione and thioredoxin), plays a key role in host-virus interactions. An excellent and comprehensive recent review is available which covers both the host and (putative) viral aspects of selenoprotein actions^287^.

The selenium content of staple cereals is strongly determined by the selenium content of soils which, prior to the use of selenium-enriched fertilisers or dietary supplements, caused regional disease outbreaks of which the iconic example is Keshan Disease; a multi-factorial syndrome whose aetiology includes an interaction between selenium deficiency and coxsackievirus B (see below)^288^.

Selenium is incorporated into the 21st amino acid, selenocysteine (where it replaces the sulphur of cysteine^287^). Gene mapping has identified 25 human selenoproteins of which 5 are glutathione reductases and 3 are thioredoxin reductases critical to the regeneration of anti-oxidant potential^287^. Activity of these enzymes is reduced in selenium deficiency. Whilst acknowledging that host-viral interactions can be modulated by both pro-oxidant and anti-oxidant factors, it is clear that anti-oxidants are key players. In this respect there are overlaps between the actions of selenium and vitamins C and E summarised elsewhere in this review.

The example of Keshan Disease provides a fascinating example of human, viral, dietary and environmental interactions with strong resonance with the emergence of SARS-CoV-2. Named after the Keshan region of China notable for selenium deficient soils, Keshan is a serious multisystem disorder affecting children and women of reproductive age^269^. A key feature is a congestive cardiomyopathy that has been linked to coxsackievirus B and can be modelled in mice. Inspired by prior studies in China^289^, Beck and colleagues passaged a benign variant of coxsackievirus B3 through selenium deficient and selenium replete mice^290,291^. The viral genome mutated in the deficient animals undergoing 6 nucleotide changes^292^, leading to myopathy and death^290,291^. Most critically, when the virus from the deficient mice was then passaged through healthy selenium-replete mice it retained its pathogenicity and caused the cardiomyopathy^290,291^. Although SARS-CoV-2 appears to be mutating slowly these studies contain a general warning that circulating viruses may be more likely to mutate to give highly-pathogenic strains with pandemic potential in nutritionally deficient populations.

A meta-analyses of almost 2 million participants in 41 randomised trials has confirmed that selenium supplementation is highly protective against Keshan disease (OR 0.14; CI 0.012-0.016)^293^. Programmes of selenium enhancement in crops and direct supplementation of the population have largely eliminated Keshan disease from the Keshan district, though it remains prevalent in neighbouring regions including Tibet and North Korea.

Beck and her team extended these studies to include the influenza A (H3N2) virus strain^268,294^. Using a similar experimental model they showed viral stability in selenium replete mice and high rates of mutation with downstream pathology in selenium deficient mice^268,294^. As with coxsackievirus the mutated strains retained their pathogenicity when re-passaged through healthy well-nourished mice^10^. Mechanisms by which selenium deficiency affect the host response to the virus were also described^295–297^. There has been little substantive new research activity in the field of selenium and viral infections by Beck’s group or others for the past decade.

### Systematic review

From a total of 12 papers returned, 4 were taken to full text screen and 2 papers were identified for full screen. One of these listed selenium as part of a COVID-19 treatment protocol but listed no results. Zhang and Liu report a general systematic review of nutrition and coronaviruses but contained no new information not already summarized above^298^.

The search of pre-print servers yielded four papers of which two were excluded. Of the remaining papers one was a systematic review^299^ and the other screened 12 organoselenium structural analogues of the antioxidant drug ebselen for inhibition of the SARS-CoV-2 papain-like protease critical to viral replication^300^. Four possible drug targets were identified.

### Clinical trials

Prior non-COVID-19 trials have investigated the impact of selenium supplementation in critically-ill patients in ICU (for a range of conditions not including ARDS). No fewer than nine meta-analyses have been performed with slightly different inclusion and grading criteria^301–309^. These analyses mostly agree that intravenous sodium selenite might yield a significant improvement in short-term mortality (meta-analysed ORs between 0.82 and 0.98), but in the latest Cochrane analysis the evidence was judged to be of very low quality due to potential to bias^302^. There was no effect on longer-term (28 or 90 day) mortality. Surprisingly, in the light of the robust animal data, there have been almost no trials of selenium and influenza or other respiratory infections. A randomised trial in 25 geriatric centres in France reported a tendency toward slightly fewer respiratory infections in patients receiving zinc and selenium, and better responses to the A/Beijing/32/92(H3N2) component of a multivalent vaccine^310^. A smaller study of a selenium-containing micronutrient supplement in English nursing homes found no effect on antibody titres after influenza vaccination^311^. In a small randomised trial, Ivory *et al*.^312^ reported no effect on mucosal influenza antibody responses to vaccination and both positive and negative effects on cellular immunity. Another small study reported that marginally deficient adults given selenium supplements had faster elimination of vaccine strains of poliovirus and fewer mutations in viral product extracted from faeces^313^.

The search of trial registers yielded two listed trials. In one of these small doses of selenium are included in the control arm. The other was a very small Phase 4 trial in which low dose selenium forms part of an antioxidant cocktail administered to both arms. There were no listed trials of intravenous selenium, suggesting that the null or very marginal results from previous trials in ICU patients have discouraged further endeavours.

## 14. Zinc

### Landscape review

Zinc is an essential trace element crucial for growth, development and the maintenance of immune function^314^. It is the second most abundant trace metal in the human body after iron, and an essential component of protein structure and function^314^. The global prevalence of zinc deficiency is estimated to range from 17-20%, with the vast majority occurring in low- and middle-income countries in Africa and Asia^315^. Zinc deficiency is also common in sub-groups of the population, including the elderly, vegans/vegetarians, and individuals with chronic disease such as liver cirrhosis or inflammatory bowel disease^314,316,317^.

Zinc is required for a wide variety of immune functions^318^ and those deficient in zinc, particularly children, are prone to increased diarrhoeal and respiratory infections. Zinc supplementation has been shown to significantly reduce the frequency and severity of both infections in children^319^, although such findings are not universal (e.g. Howie *et al*.^320^) and a recent systematic review and meta-analysis found no evidence that adjunctive zinc treatment improves recovery from pneumonia in children in low- and middle-income countries^321^. Similar to vitamin C, zinc supplementation has also been suggested as a potential remedy for the treatment of the common cold (rhinovirus infection); a meta-analysis of 3 trials reporting on 199 patients supports a faster recovery time^322^ although the small sample size (N=199) of included studies warrants caution.

At the molecular level, zinc is an essential component of protein structure and function and is a structural constituent of ∼750 zinc-finger transcription factors, enabling gene transcription^314,323^. It is also a catalytic component of approximately 2000 enzymes^324^. The role of zinc homeostasis in antibacterial immune responses is well-documented; binding and sequestering extracellular zinc (and calcium) can prevent bacterial and fungal overgrowth^325^ while toxic endosomal zinc accumulation can inhibit intracellular Mycobacterium growth in macrophages^326^. For viral infections, however, these mechanisms are less well described although a number of new hypotheses are now being suggested^327^.

The SARS-CoV-2 pandemic has resulted in a global search for suitable antiviral and immunomodulatory candidates. Attracting global attention at the start of the pandemic was the potential use of oral chloroquine (CQ) and hydroxychloroquine (HQ), prescription drugs normally used for the treatment of malaria. Emerging trial evidence, however, does not support the use of either CQ or HQ as a treatment option for the disease^328–330^. Of relevance to the current review is the finding that CQ has characteristics of a zinc ionophore and specifically targets extracellular zinc to intracellular lysosomes^331^. This has led to an interest in zinc as a potential target for anti-viral therapies, most notably in combination with CQ/HQ in clinical trials for the prevention or treatment of SARS-CoV-2^332^.

### Systematic review

From a total of 69 papers returned (after removal of eight duplicates), six were taken to full text screen. On full screen five papers were rejected as ineligible and one review paper, although ineligible for this review as it included no new data presented, highlighted the potential synergistic action of zinc and CQ in patients with SARS-CoV-2^333^.

A review of preprint listings returned 10 potentially relevant papers. Five of these were duplicates (already identified via PubMed or EMBASE). Four were found to be review articles, with no novel data specific to COVID-19 disease susceptibility or progression. Only a single paper was eligible for inclusion, a retrospective observational study comparing hospital outcomes (New York, USA) among patients who received HQ and azithromycin plus zinc versus HQ and azithromycin alone^334^. Using data from 932 patients admitted over a one-month period (March-April 2020) the authors found that addition of zinc sulphate did not impact the length of hospitalization, duration of ventilation, or ICU duration. In univariate analyses, zinc sulphate increased the frequency of patients being discharged home, and decreased the need for ventilation, admission to the ICU, and mortality or transfer to hospice for patients who were never admitted to the ICU. After adjusting for the time at which zinc sulphate was added to the protocol, an increased frequency of being discharged home (OR 1.53, 95% CI 1.12-2.09), and a reduction in mortality or transfer to hospice remained significant (OR 0.449, 95% CI 0.271-0.744). These data provide initial *in vivo* evidence that zinc sulphate may play a role in therapeutic management for COVID-19.

### Clinical trials

A screen of registered trials revealed 16 studies of potential relevance. On review, three were removed; one was an observational case-control study and in a further two studies zinc was not included in any of the experimental arms. Of the remaining 13 trials, only a single trial (USA, n=520) is designed to fully assess the impact of zinc, in a four-arm trial of outpatients who test positive for SARS-CoV-2 and comparing vitamin C (8000mg/d) vs zinc (50mg/d) vs vitamin C + zinc (doses as before) vs standard of care (NCT04342728). In a further treatment trial among SARS-CoV-2 patients in Senegal (n=384), zinc (20mg/d) is being used as the control arm in a trial of HQ plus azithromycin (two arms with differing dosing regimens) (PACTR202005622389003). In four trials, zinc (at doses ranging from 15 to 250mg/d) is being administered in combination with other antiviral drugs including HQ, HC and azithromycin, HC and doxycycline or favipiravir. In the remaining seven trials, zinc is being provided in combination with single or multiple other micronutrients, including vitamin C, and vitamin B12 and, therefore, the potential therapeutic benefits of zinc as a single micronutrient cannot be established.

## 15. Antioxidants

### Landscape review

During severe COVID-19, the SARS-CoV2 virus can trigger a strong host immune response. This can then result in the production of high levels of free radicals by both macrophages and neutrophils and the induction of severe oxidative stress^335^. Oxidative stress causes protein and lipid oxidation which then further activates and amplifies the immune response creating a self-amplifying loop which can result in extensive tissue damage^336^.

Oxidative stress is currently thought to be a major cause of the pathophysiology of severe COVID-19 infections and has previously been implicated as a mediator in acute respiratory distress syndrome^337^. The level of oxidative stress may indeed determine the intensity of the organ damage seen during severe COVID-19 specifically to endothelial, pulmonary, cardiac and immune cells^338^. In addition, increased levels of oxidative stress pre-exist in individuals with co-morbidities such as obesity, diabetes and cardiovascular disease, and may play a role in increasing the risk of severe COVID-19 in these groups^339^.

Antioxidants decrease oxidative stress and can be broadly divided into four groups: (1) Endogenous antioxidants which include molecules (e.g. glutathione, uric acid and transferrin), vitamins (such as Vitamin A, C, and E) and enzymatic co-factors (e.g. selenium and zinc) synthesized by the human body; (2) Dietary antioxidant molecules and vitamins found in food (e.g. fruit, vegetables, green tea, olive oil and red wine); (3) Nutritional supplement antioxidants which include supplements that contain increased doses of dietary antioxidants (e.g. vitamin C or quercetin tablets), molecules from medicinal plants (e.g. molecules found in traditional Chinese medicine), and (4) Synthetic molecules or drugs with known antioxidant activities (e.g. N-acetyl cysteine and metformin).

There is an abundance of epidemiological and *in vitro* evidence to suggest that levels of endogenous antioxidants and increased consumption of dietary antioxidants may decrease inflammation and oxidative stress^213^, particularly in patients with cardiovascular disease^340^.

However, there is a lack of clinical evidence that consuming antioxidants from dietary sources or giving acute doses of naturally occurring antioxidants has direct long-term clinical benefits in the treatment of chronic conditions or acute viral infections^341^. Some relevant evidence exists for the clinical utility of a synthetic antioxidant, N-acetyl cysteine, which is also an FDA-approved drug for the treatment of paracetamol toxicity. N-acetyl cysteine has been shown to have some modest benefit in ARDS^342^ and there is limited evidence that it improves clinical outcomes in several viral diseases including HIV^343^, hepatitis A^344^, H1N1 influenza^345^, dengue^346–348^, and rotavirus infection^349^.

### Systematic review

From a total of 212 papers returned, 44 were taken to full text screen. Nineteen papers were commentaries or non-systematic reviews. In no case was there any new data related to antioxidants as a clinical therapy for COVID-19. Note that information on COVID-19 and vitamins A, C and E as well as selenium and zinc have been reviewed in separate sections of this manuscript and those papers were not included here.

The search of preprint servers yielded six relevant papers all of which were accessed for full review. All were commentaries or editorials. None contained any new data on antioxidants as a treatment or preventative therapy for COVID-19.

### Clinical trials

The search of clinical trials registers yielded eight entries involving antioxidants (that were not vitamins A, C or E) (Supplementary Material 4). Of the eight trials, three involved dietary supplements containing a mixture of antioxidants and other molecules. The remaining five are testing the following molecules: Reservatrol, Silymarin, Quercetin, N-acetyl cysteine and melatonin. These trials target participants at various stages of SARS-CoV2 infection.

## 16. Nutritional Support

### Landscape Review

Evidence on best practice for nutritional support for patients with COVID-19 is currently lacking^350^. In those infected, 80% have a mild condition (not requiring hospitalisation) whilst 20% require inpatient care and 5% will require intensive care^351,352^. In the 80% with mild disease there is a growing body of evidence that the course of illness may take several weeks and in some cases many months for recovery and have multiple complications along the way^353^.

In those admitted to hospital, nutritional support guidelines and advice are generally based on evidence drawn from treatment of viral pneumonia, sepsis and ARDS. Specific evidence in relation to COVID-19 is not available as yet, but a pragmatic approach and “doing what we know, and doing it well” has been adopted in most settings. There is a huge wealth of literature on nutritional support in critically ill patients^354^, which is beyond the scope of this review, but we will briefly discuss consensus on best practice.

Nutritional support during an acute illness has long been recognised as an important component to care^355^. In an acute, severe illness there is a high risk of catabolism and the resulting malnutrition and sarcopenia can impact both on mortality and morbidity^354,356^.

Recommended nutritional support varies in mild, severe and critical disease but there are overarching considerations which can be divided into patient factors, healthcare staffing factors and system factors^357^. Patient factors overlap in all disease states. There may be the need for special nutritional intervention in mild disease especially in those with pre-existing conditions such as diabetes, heart failure and other cardiac or chronic diseases. These may be exacerbated by an acute viral illness, especially if diarrhoea, vomiting or anorexia are present. A study from a rehabilitation centre in Italy, focused on patients once they were past the acute phase of their illness, found 45% of COVID-19 infected patients were at risk of malnutrition^358^. At the peak of cases, when healthcare systems have the potential to be overrun, the staffing shortages and other demands would make this easy to neglect to the detriment of the patients.

Patients with severe disease are usually admitted to hospital. There is consensus that all patients admitted with COVID-19 should have their nutritional status assessed^56,359^. There are a number of important and practical considerations that affect nutritional care:

- Risk of hypoxia on removal of oxygen delivery device (mask or non-invasive ventilation) to eat and drink.
- Ability to remove oxygen delivery device independently to eat and drink.
- Ease of access to food and drink.
- Air leakage with non-invasive ventilation (NIV) mask due to nasogastric (NG) tube.

These factors, along with isolation of COVID-19 patients in single rooms, limited visits by healthcare workers due to the need to conserve PPE and reduce risk of transmission, and limited visits by family or friends, mean there is a real danger of malnutrition and dehydration^359^.

A solution to this is the adoption of an early nutritional supplementation program as detailed in the study by Caccialanza *et al*^56^. In this feeding protocol all patients were screened at admission using a simplified nutritional risk score and due to a high number of patients being unable to meet their nutritional needs on a normal diet, all patients were started on whey proteins (20g/day) and multivitamins, multiminerals and trace elements supplement. Those at nutritional risk were then commenced on 2-3 bottles of Oral Nutritional Supplements (ONS) and escalated to parenteral nutrition (PN) should they be unable to tolerate oral intake.

Another solution adopted in the UK is the “Every Contact Counts” model, where patients are offered food and drink at every encounter with health professionals^359^.

Both the consensus statement by nursing practitioners in China and the ESPEN expert statement agree on the following steps^58,360^:

- Early screening for risk of malnutrition
- Individualized nutritional plans
- Oral Nutritional Supplements to be used
- Parenteral nutrition should be initiated within 3 days should enteral nutrition (EN) not meet nutritional requirements
- Ongoing monitoring of nutritional status

ESPEN give the following additional details:

- Aim for 30 kcal/kg/day to meet energy needs (may need to be adjusted in certain populations)
- 1g/protein/day (may need to be adjusted in certain populations)
- Fat: carbohydrate ratio in non-ventilated patients 30:70

Whichever approach is taken, prevention of inpatient malnutrition and its associated complications must be considered an essential component to clinical care and requires monitoring throughout the illness.

The ESPEN guidelines on nutritional support of patients admitted to ICU and the document specifying treatment in those with COVID-19 are thorough and comprehensive^58,354^. These guidelines as well as the guidelines from the American Society for Enteral and Parenteral Nutrition are based on evidence of feeding in critically ill patients and expert opinion on how that can be applied to COVID-19^350^.

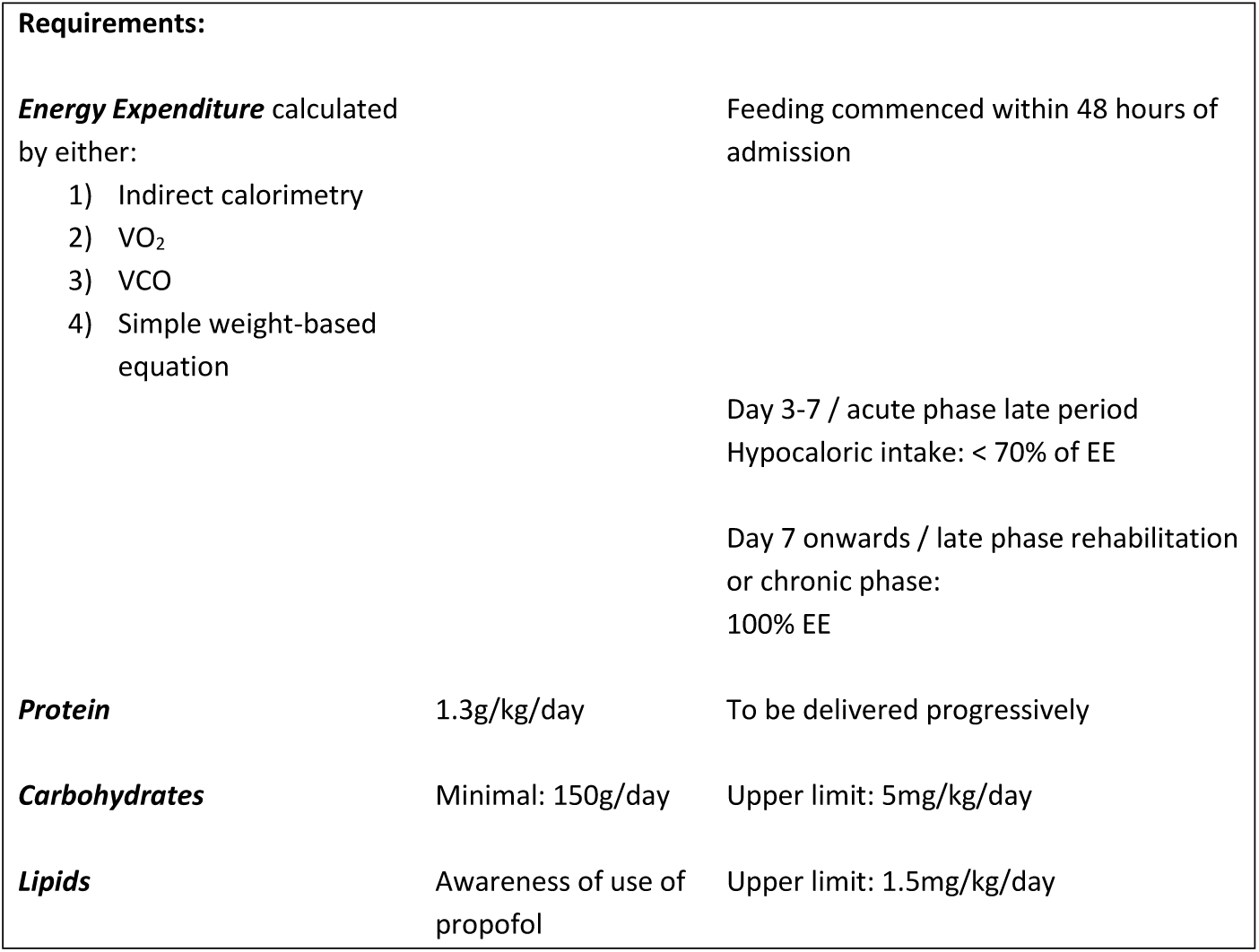

COVID-19 patients in ICU have a few special considerations relating to treatment and nutrition. For example, many patients required proning during the course of their ventilation and there is consensus in all guidelines on EN feeding being safe to continue as long as awareness of complications with NG placement and specified steps to minimize this were taken. Depending on the severity of lung injury and its availability, there can be many patients requiring extracorporeal membrane oxygenation (ECMO) and there was consensus in all guidelines that EN feeding can be started at trophic or hypocaloric levels. Use of PN differed, with American guidelines advocating early implementation and European and Chinese guidelines taking a more cautious case-by-case approach. Finally nutritional support may well be needed post ICU discharge with high rates of dysphagia being reported^358,361^.

### Systematic review

Our systematic search yielded 17 papers for full review, none of which met the criteria for inclusion. Of the pre-prints, 15 were reviewed and none met the full criteria.

### Clinical trials

Sixteen clinical trials were identified through our search and two were relevant to nutritional support (Supplementary Material 4). One has not yet started recruitment and aims to describe nutritional consequences of COVID-19 in patients discharged from hospital (based in France). The other aims to validate the use of a nutrition scoring tool “NUTRIC” in Chinese ICU patients diagnosed with COVID-19, results pending.

## 17. Discussion

As the pandemic continues to evolve at rapid pace, so does our understanding of the epidemiology and underlying mechanisms of the SARS-CoV-2 virus. However, despite the wealth of literature being published, the evidence directly linking nutritional status to the risk and progression of COVID-19 is still sparse. In Figure 2 we summarise the key themes emerging from our landscape and systematic reviews.

**Figure 2:**
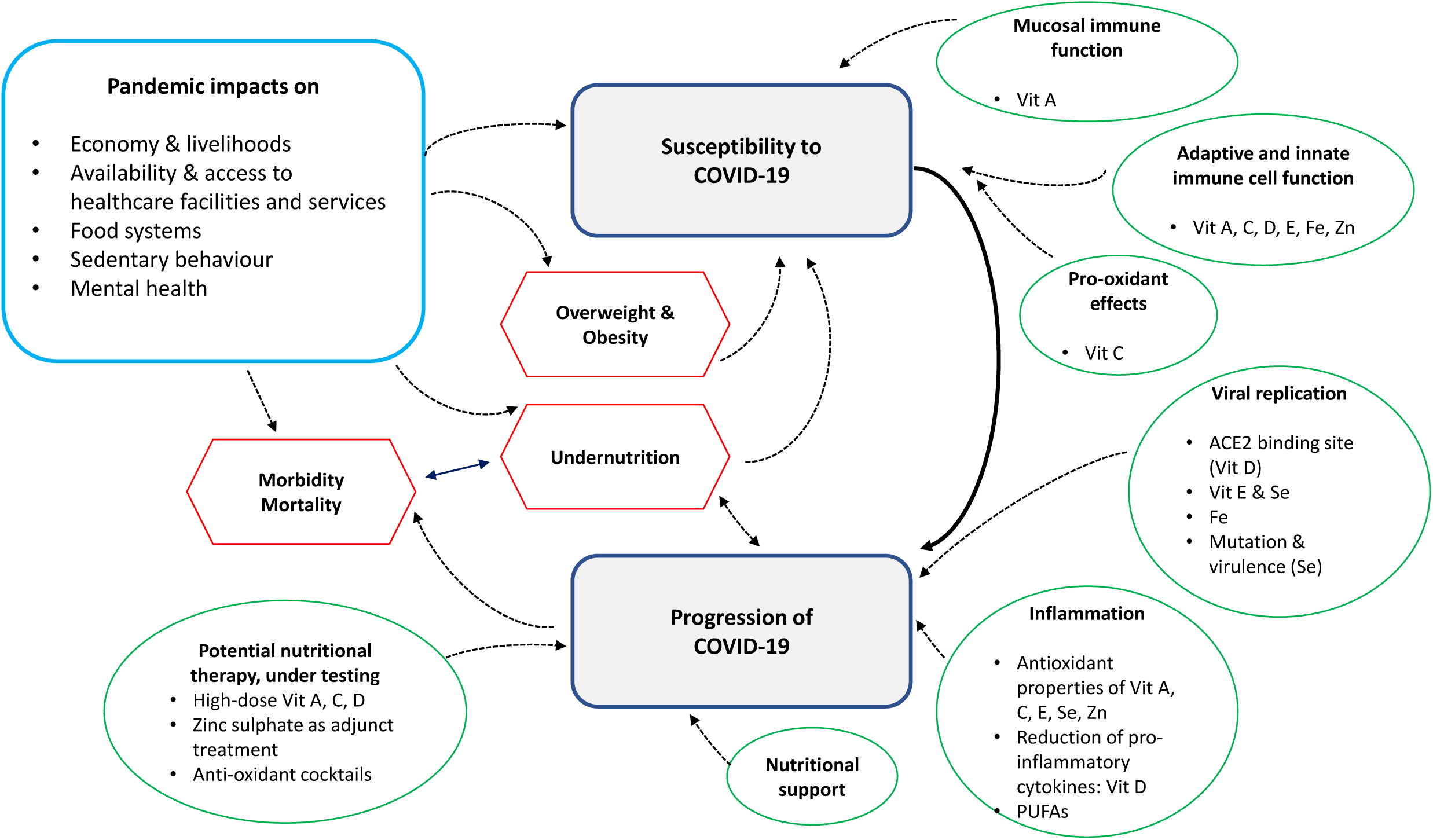
Overview diagram showing key concepts drawn from narrative synthesis.

Nutritional status has the potential to influence susceptibility to the risk of COVID-19 through its integral role in immune function. For example, above we have covered some of the ways micronutrients support mucosal immune function (vitamin A), epithelial tissue integrity (vitamins A, C and D), enhancing the function of certain adaptive and innate immune cells (vitamins A, C, D, E, iron, zinc and PUFAs) and potential pro-oxidant effects (vitamin C). Undernutrition, overweight, obesity and type-II diabetes are all associated with impaired immunity, through independent (though as yet not clearly defined) mechanisms as well as through the effects of concurrent micronutrient deficiencies. The various presentations of overnutrition have been the most frequently documented nutrition-related co-morbidities amongst patients admitted to hospital with COVID-19 to date. However, many markers of micronutrient deficiency are not routinely measured on hospital admission. Furthermore, at the time of writing, the pandemic is still penetrating LMICs, where the burden of undernutrition is higher. We therefore anticipate further evidence on the potential impact of undernutrition on COVID-19 susceptibility to be generated soon.

The influence of nutrition on immune function can also affect the progression of viral infections, with implications for the length, severity and final outcomes of disease episodes. From our landscape reviews we only have limited insight from other viral diseases as to how nutritional supplementation may potentially influence outcomes. For example, although there is strong evidence of an association between vitamin A supplementation and reduced outbreaks of measles, there is insufficient evidence regarding the association with Ebola outcomes. For vitamin C there is some positive, but inconsistent, evidence regarding supplementation and the prevention of pneumonia, but very limited evidence describing an effect of supplementation on overall mortality reduction. For vitamin D we have mixed evidence describing the influence of supplementation on both the risk and severity of acute respiratory infections. For the minerals, we have documented evidence of an association between iron deficiency and increased risk of impaired lung function in hypoxic conditions, and literature describing the association between zinc supplementation and reduction of diarrhoea and respiratory infections. It is important to note, however, that not all evidence of nutritional supplementation points to positive, or null, outcomes. For example, we have described how there is some evidence linking vitamin E supplementation to the worsening of respiratory infections. Furthermore, some studies have found evidence of associations between iron supplementation or elevated iron status with increased risk of malaria, bacterial infections, HIV-1 progression, and certain respiratory infections.

However, when it comes to COVID-19 explicitly, our ability to draw conclusions between nutritional status and disease progression is limited by the current lack of high-quality data. We have documented some observational studies describing an association between lower vitamin D status and increased COVID-19 infection. We noted that a single observational study suggested treatment with zinc sulphate showed signs of reduction in mortality and increased discharge from hospital to home in patients treated with hydroxychloroquine and azithromycin. However, more recent findings from the Recovery trial find no beneficial effect of HQ in the absence of zinc^362^. We also summarised some observational studies that described how patients presenting with malnutrition on hospital admission (both under- and over-nutrition) have increased risk of mortality from COVID-19. With studies on undernutrition in particular, it is not easy to distinguish between the effect of pre-existing undernutrition on immune function and increased disease severity, and the subsequent nutritional impact of prolonged inflammatory states and intensive care admission through impaired appetite and dysregulated metabolism.

The literature has, however, highlighted some hypotheses regarding mechanisms through which nutrition could modulate disease severity and progression. Particularly relevant to COVID-19 is the role anti-oxidants may play in reducing the impact of the cytokine storm during the acute phase of the infection. This has to be carefully balanced against not overly dampening the immune response during other phases of the illness, as described in detail in Iddir *et al*.^213^. Of the micronutrients covered in our review, vitamins A, C, E, and certain dietary polyphenols have potentially important roles in quenching free radicals through their anti-oxidant properties, alongside zinc and selenium in their coenzyme roles. Synthetic anti-oxidants can be produced and are being tested for effectiveness in mitigating the damage from the cytokine storm, and it is not yet clear to what extent dietary components will play a synergistic role.

Micronutrients may help slow down processes vital for viral replication. For example, we have described how vitamin D may influence the expression of ACE2, implicated in SARS-CoV-2 binding. Animal studies have shown, tentatively, how deficiencies in selenium and vitamin E may increase viral replication as well as enhancing virulence and mutation rates.

To date, the role of nutritional support in the clinical management of severe COVID-19 cases is based on knowledge from successful protocols used in other viral infections and, more generally, in recovery from intensive care. There are, however, some new treatment regimens being tested. Treatments comprising combinations of various antioxidants are currently being investigated in the early stages of intervention trials. It is not be possible to separate out the effects of individual micronutrients in these treatments. Higher doses of vitamins A, C, and D are also being trialled, some intra-venously, but there is limited prior evidence to suggest they will be successful and many trials do not seem to take account of normal physiological thresholds. For the minerals, the potential role of iron chelation in reducing iron-induced lung toxicity is being considered. Zinc features mainly as an adjunct therapy alongside chloroquine and hydroxychloroquine interventions, although interest is growing in its potential as an intervention in its own right. Nutritional supplementation will require careful consideration of the extent to which the suggested micronutrients can be utilised, especially during acute inflammation and the related states of anaemia of inflammation. It is likely a period of stabilisation to bring down inflammation will be essential before any positive effects from micronutrient supplementation can be seen^156,363^.

In this review we have focussed on the direct relationship between nutritional status and risk of infection and progression of COVID-19. This is an important but incomplete part of the vicious cycle of nutritional status, immune response and infection. Beyond the scope of this review, but integral to the overall picture, are the impacts the pandemic has on livelihoods and health, that are inextricably linked to nutritional status and therefore overall morbidity and mortality. We know from the Ebola outbreak in West Africa during 2013-16 that disruption to the health system brought about excess mortality equal to, if not greater than, direct deaths from the infection itself^364^. The disruption from COVID-19 to food systems, the economy and health infrastructure means that nutritional status of the most vulnerable will be enormously impacted. Headey *et al*. summarise recent estimates from modelling, suggesting that an additional 140 million people are expected to fall into extreme poverty due to the pandemic in 2020 alone, with a doubling of people facing food insecurity (estimated at 265 million)^8^. An estimated 14.3% increase in wasting prevalence in children under 5 will equate to an additional 6.7 million children wasted compared to estimates without COVID-19^8^. Furthermore, the increase in numbers of people facing acute nutritional vulnerability will be compounded by the reduction in health services offered to the population during the pandemic. Roberton *et al*. modelled scenarios estimating impacts of different levels of disruption to availability of health workers and supplies, and on demand and access to health services. Even in the best case scenario they estimated the additional prevalence of acute malnutrition and reduced coverage of health services would result in an additional quarter of a million child deaths in the next 6 months^365^.

Many consortia have highlighted the urgency of tackling the immense impact of the pandemic on nutrition and health outlined above. Recommendations point both to nutrition-specific strategies, such as prevention and treatment of wasting, vitamin A supplementation, and breast-feeding support;^366^ and to nutrition-sensitive strategies, such as strengthening the food-supply chain, providing safety net programmes, implementing community-led sanitation initiatives, improving female empowerment, and ensuring access to healthcare^9^.

### Strengths and limitations of the review

Our review provides a synthesis of information to complement other existing comprehensive reviews^207,213^. However, to our knowledge ours is the most detailed systematic search to date, bringing together 13 separate systematic reviews. Our inclusion of material from pre-print servers and trial registries adds to the breadth of information we have been able to include.

The pandemic is evolving rapidly and new evidence has likely surfaced since our search dates. Whilst the collation of 13 reviews in this article provided breadth, we were unable to ensure all searches took place exactly synchronously. We did not perform a risk of bias assessment of the included literature, and it is important to note that pre-prints are not peer-reviewed. Our inclusion criteria of literature written in English may have missed some pertinent information in other journals. We necessarily had to limit our scope to the most important nutrition-related conditions and micronutrients of interest. However, this is incomplete, and other potentially relevant areas of interest include the role of macronutrient intake, gut microbiota, dietary fibre, B vitamins, other minerals, phytochemicals, and carotenoids. These are covered in other narrative reviews^210,213^. Furthermore, we were unable to comprehensively cover all the additional factors that can influence the relationship between nutrition, immunity and disease progression. Interpretation of the included literature is necessarily restricted to the context of the original studies, and a wide range of factors (some measured, many not measured) preclude extrapolation to the wider population. Such considerations should include genetic polymorphisms and their frequency and impact in different populations, haemoglobinopathies, the environment (e.g. soil type, latitude), age, sex, access to healthcare, and other underlying economic and political factors determining nutritional vulnerability. Finally, there is always a degree of uncertainty and risk when extrapolating from one infection to another, especially when age profiles of the affected population vary. We find that much of the previous literature on micronutrient deficiencies and viral infection focus on the younger population, whereas SARS-CoV-2 is predominantly affecting older people.

## Conclusion

Our review of the current literature highlights a range of mechanistic and observational evidence to highlight the role nutrition can play in susceptibility and progression of COVID-19. Prior knowledge of interactions between nutrition and other viral diseases can help inform hypotheses relevant to COVID-19. However, the literature taken from other viral diseases is far from consistent, and studies taken in isolation can be a source of rumours and ill-advised quick-fixes surrounding COVID-19 prevention and cure. There is limited evidence to date that high-dose supplements of micronutrients will either prevent disease or speed up treatment. Attempting to ensure people have an adequate dietary intake is critical. However, we believe the focus should be on ways to promote a balanced diet and reduce the infective burden rather than reliance on high-dose supplementation, until more concrete evidence from clinical trials suggests otherwise. Whilst the quantity of literature on these topics is increasing daily, this does not necessarily correspond to an increase in high-quality evidence. Reviews such as ours will continually need updating to allow for a balanced view of the available data in order to counter unjustified nutrition-related claims. To date there is no evidence supporting adoption of novel nutritional therapies, although results of clinical trials are eagerly awaited. Given the known impacts of all forms of malnutrition on the immune system, public health strategies to reduce micronutrient deficiencies, undernutrition and over-nutrition remain of critical importance, drawing on the numerous lessons learnt from other viral diseases.

## Data Availability

All data contributing to the narrative synthesis is made available within the article and supplementary materials.

## Funding acknowledgements

ZA and PS are supported by the Wellcome Trust Our Planet Our Health Programme (FACE-Africa grant number: 216021/Z/19/Z). MJ is supported by Wellcome Trust grant (ref 216451/Z/19/Z). HD, AEA and MT are supported by UK Medical Research Council (MRC Human Immunology Unit core funding to H.D., award no. MC_UU_12010/10). SEM is supported by The Wellcome Trust (ref 220225/Z/20/Z) and the Medical Research Council (UK) (ref MR/P012019/1). AMP, CC and MJS are jointly funded by the UK Medical Research Council and the Department for International Development (DFID) under the MRC/DFID Concordat agreement (MRC Programme MC-A760-5QX00). KSJ is supported by the National Institute for Health Research (NIHR) Cambridge Biomedical Research Centre (IS-BRC-1215-20014). The NIHR Cambridge Biomedical Research Centre is a partnership between Cambridge University Hospitals NHS Foundation Trust and the University of Cambridge, funded by the NIHR. The views expressed are those of the authors and not necessarily those of the NHS, the NIHR or the Department of Health and Social Care. All other authors received no specific funding for this work.

## Competing interests

The authors have declared that no competing interests exist.

## Supplementary Material 1: Search Strategy

This review looks at how malnutrition in all its forms (undernutrition, micronutrient deficiencies and overnutrition) may influence both susceptibility to, and progression of, COVID-19. We synthesise information on the following 13 nutrition-related components and their potential interactions with COVID-19:

i. Protein-energy malnutrition
ii. Overweight, obesity and diabetes
iii. Anaemia
iv. Iron
v. Vitamin A
vi. Vitamin C
vii. Vitamin D
viii. Vitamin E
ix. Poly-Unsaturated Fatty Acids
x. Selenium
xi. Zinc
xii. Anti-oxidants
xiii. Nutritional support

Each section follows the following structure:

### 1. Landscape review of other pertinent evidence

This section does not require a systematic search. Coverage is limited to: a) very brief description of nutrient/condition vis-à-vis infection/immunity; b) evidence of any role in other viral infections (especially of respiratory tract); c) possible mechanisms; d) possible utility in treatment.

### 2. Systematic review of published literature and pre-prints

a. PUBMED – see example search string in **Supplementary Material 2**
b. EMBASE - see example search string below **Supplementary Material 2**
c. Pre-print servers: see search terms in **Supplementary Material 2**
  i. WHO Global literature on coronavirus disease: https://search.bvsalud.org/global-literature-on-novel-coronavirus-2019-ncov/
  ii. The Lancet COVID-19 Resource Centre: https://www.thelancet.com/coronavirus
  iii. The JAMA network Coronavirus Resource site: https://jamanetwork.com/collections/46099/coronavirus-covid19
  iv. The New England Journal of Medicine Coronavirus Resource site: https://www.nejm.org/coronavirus
  v. The bioRxiv preprint server: https://www.biorxiv.org
  vi. The medRxiv preprint server: https://www.medrxiv.org
  vii. The ChinaXiv preprint server: http://chinaxiv.org/home.htm
  viii. The ChemRxiv preprint server: https://chemrxiv.org/
  ix. The Preprints server: https://www.preprints.org
  x. The Research Square preprint site: https://www.researchsquare.com
  xi. The LitCovid hub: https://www.ncbi.nlm.nih.gov/research/coronavirus/
  xii. The WHO Global research database: https://www.who.int/emergencies/diseases/novel-coronavirus-2019/global-research-on-novel-coronavirus-2019-ncov
  xiii. The Cell Press Coronavirus Resource Hub: https://www.cell.com/2019-nCOV
  xiv. The Nature Research Coronavirus collection: https://www.nature.com/collections/hajgidghjb
  xv. Science Coronavirus collection: https://www.sciencemag.org/collections/coronavirus
  xvi. The COVID-19 Primer: https://covid19primer.com/dashboard

#### Inclusion criteria

- Human studies
- **PubMed & EMBASE:** related to COVID-19, MERS-CoV or SARS-CoV AND disease susceptibility / progression AND nutrient exposure of interest.
- **Pre-print servers:** related to COVID-19 AND disease susceptibility / progression AND nutrient exposure of interest.
- All original studies of any design
- Systematic reviews (to check bibliography)
- Published in English language

#### Exclusion criteria

- Comments, letters, opinions, non-systematic reviews

### 3. Systematic review of the following clinical trial registers

a. ClinicalTrials.gov: https://clinicaltrials.gov/
b. ISRCTN Registry: https://www.isrctn.com/
c. EU Clinical Trials Register: https://www.clinicaltrialsregister.eu/
d. Pan African Clinical Trials Registry: https://pactr.samrc.ac.za/
e. India Clinical Trials Registry: http://ctri.nic.in/Clinicaltrials/login.php
f. Chinese Clinical Trial Registry: http://www.chictr.org.cn/enIndex.aspx

**Inclusion criteria:** trials related to COVID-19 AND nutrient exposure of interest, human trials, protocols in English language.

See **Supplementary Material 2** for simplified search terms.

## Supplementary Material 2: Search terms

### Search terms for PubMed and EMBASE databases

|  | Concept 1 AND | (Concept 2 OR | Concept 3) | AND Concept 4 |
| --- | --- | --- | --- | --- |
| Key concepts | Coronavirus **AND** | (Disease susceptibility **OR** | Disease progression) | **AND**<br>Terms specific to sub-section |
| Free text terms | coronavir* OR<br>"coronavirus infections" OR<br>covid* OR ncov* OR 2019-ncov<br>OR 2019ncov OR "2019-novel<br>CoV" OR HCoV* OR cov2 OR<br>"cov 2" OR OC43 OR NL63 OR<br>229E OR HKU1 OR "sars<br>coronavirus 2" OR "sars-like<br>coronavirus" OR<br>"Severe Acute Respiratory<br>Syndrome" OR SARS OR sars-cov* OR sarscov* OR<br>"Middle East Respiratory<br>Syndrome" OR MERS OR<br>MERS-CoV | "adaptive immunity" OR<br>"acquired immunity" OR<br>"innate immunity" OR<br>"cell-mediated immunity" OR<br>"humoral immunity" OR<br>"antibody formation" OR<br>immunosuppression OR<br>immunodepression OR<br>"immunity impairment" OR<br>"immune dysfunction" OR<br>"lymphocyte function" OR<br>"macrophage activity" OR<br>"oxidative stress" OR<br>"host defence" OR<br>"immune response" OR<br>inflammation OR<br>"immune pathology" OR<br>immunopathology OR<br>"Macrophage activation<br>syndrome" OR<br>"MAS" OR<br>"cytokine storm" | "viral load" OR<br>"viral pathogen*" OR<br>"viral replication" OR<br>"viral mutation" OR<br>"viral transmission" OR<br>"acute respiratory distress OR<br>syndrome" OR<br>"ARDS" OR<br>"hemophagocytic<br>lymphohistiocytosis" OR<br>"HLH" OR<br>pneumonia OR bronchitis OR<br>bronchiolitis OR "asthma<br>exacerbation*" OR seizure*<br>OR diarrhoea OR diarrhea OR<br>"acute gastroenteritis" or<br>dehydration or "electrolyte<br>imbalance" OR "renal failure"<br>OR "kidney failure" OR "multi-organ failure*" OR "multiple<br>organ failure*" OR<br>encephalomyelitis OR<br>"Kawasaki disease" OR<br>"Kawasaki syndrome" OR<br>"Mucocutaneous Lymph Node<br>Syndrome" OR coagulopathy<br>OR death OR mortality | e.g. Vit C<br>Ascorbic acid |
| **Controlled vocabulary terms / Subject terms**<br>MeSH terms | "Coronavirus Infections"[Mesh] OR<br>"Coronavirus"[Mesh] OR<br>"COVID-19"[Supplementary Concept] OR<br>"Severe Acute Respiratory Syndrome"[Mesh] OR<br>"severe acute respiratory syndrome coronavirus 2"[Supplementary Concept] OR<br>"Middle East Respiratory Syndrome Coronavirus"[Mesh] | "Immune System Phenomena"[Mesh] OR<br>"T-Lymphocytes, Regulatory"[Mesh] OR<br>"Inflammation"[Mesh] OR<br>"Immunosuppression"[Mesh] OR<br>"Oxidative Stress"[Mesh] OR<br>"Macrophage Activation Syndrome"[Mesh] | "Viral Load"[Mesh] OR<br>"Virus Physiological Phenomena"[Mesh] OR<br>"Respiratory Distress Syndrome, Adult"[Mesh] OR<br>"Respiratory Tract Infections"[Mesh] OR<br>"Gastrointestinal Diseases"[Mesh] OR<br>"Gastroenteritis"[Mesh] OR<br>"Seizure"[Mesh] OR<br>"Diarrhea"[Mesh] OR<br>"Dehydration"[Mesh] OR<br>"Water-Electrolyte Imbalance"[Mesh] OR<br>"Kidney Failure, Chronic"[Mesh] OR<br>"Shock"[Mesh] OR<br>"Encephalomyelitis"[Mesh] OR<br>"Mucocutaneous Lymph Node Syndrome"[Mesh] OR<br>"Blood Coagulation Disorders"[Mesh] OR<br>"Mortality"[Mesh] | e.g.<br>"Ascorbic Acid"[Mesh] |
| **Controlled vocabulary terms / Subject terms**<br>Emtree terms | exp Coronavirus/<br>exp Coronaviridae infection/<br>exp severe acute respiratory syndrome/<br>exp SARS coronavirus/<br>exp Middle East respiratory syndrome coronavirus/ | exp immune system/<br>exp T lymphocyte/<br>exp inflammation/<br>exp immune deficiency/<br>exp oxidative stress/<br>exp macrophage activation/<br>exp cytokine storm/ | exp virus load/<br>exp virus transmission/<br>exp virus shedding/<br>exp virus virulence/<br>exp virus characterization/<br>exp virus immunity/<br>exp virus infection/<br>exp virus cell interaction/<br>exp virus transcription/<br>exp virus inhibition/<br>exp respiratory tract infection/ | exp ascorbic acid/ |
|  |  |  | exp gastrointestinal infection/<br>exp pneumonia/<br>exp virus pneumonia/<br>exp bronchitis/<br>exp bronchiolitis/<br>exp viral bronchiolitis/<br>exp asthma/<br>exp seizure/<br>exp diarrhea/<br>exp gastroenteritis/<br>exp viral gastroenteritis/<br>exp dehydration/<br>exp electrolyte disturbance/<br>exp kidney failure/<br>exp multiple organ failure/<br>exp encephalomyelitis/<br>exp mucocutaneous lymph<br>node syndrome/<br>exp blood clotting disorder/<br>exp mortality/ |  |

#### Vitamin C example

### Example search string in PubMed

(coronavir* OR “coronavirus infections” OR covid* OR ncov* OR “2019-ncov” OR “2019ncov” OR “2019-novel CoV” OR HCoV* OR cov2 OR “cov 2” OR OC43 OR NL63 OR 229E OR HKU1 OR “sars coronavirus 2” OR “sars-like coronavirus” OR “Severe Acute Respiratory Syndrome” OR SARS OR sars-cov* OR sarscov* OR “Middle East Respiratory Syndrome” OR MERS OR “MERS-CoV” OR “Coronavirus Infections”[Mesh] OR “Coronavirus”[Mesh] OR “COVID-19”[Supplementary Concept] OR “Severe Acute Respiratory Syndrome”[Mesh] OR “severe acute respiratory syndrome coronavirus 2”[Supplementary Concept] OR “Middle East Respiratory Syndrome Coronavirus”[Mesh])

#### AND

((“adaptive immunity” OR “acquired immunity” OR “innate immunity” OR “cell-mediated immunity” OR “humoral immunity” OR “antibody formation” OR immunosuppression OR immunodepression OR “immunity impairment” OR “immune dysfunction” OR “lymphocyte function” OR “macrophage activity” OR “oxidative stress” OR “host defence” OR “immune response” OR inflammation OR “immune pathology” OR immunopathology OR “Macrophage activation syndrome” OR “MAS” OR “cytokine storm” OR “Immune System Phenomena”[Mesh] OR “T-Lymphocytes, Regulatory”[Mesh] OR “Inflammation”[Mesh] OR “Immunosuppression”[Mesh] OR “Oxidative Stress”[Mesh] OR “Macrophage Activation Syndrome”[Mesh])

#### OR

(“viral load” OR “viral pathogen*” OR “viral replication” OR “viral mutation” OR “viral transmission” OR “acute respiratory distress syndrome” OR “ARDS” OR “hemophagocytic lymphohistiocytosis” OR “HLH” OR “pneumonia” OR “bronchitis” OR “bronchiolitis” OR “asthma exacerbation*” OR “seizure*” OR “diarrhoea” OR “diarrhea” OR “acute gastroenteritis” or “dehydration” or “electrolyte imbalance” OR “renal failure” OR “kidney failure” OR “multi-organ failure*” OR “multiple organ failure*” OR “encephalomyelitis” OR “Kawasaki disease” OR “Kawasaki syndrome” OR “Mucocutaneous Lymph Node Syndrome” OR “coagulopathy” OR “death” OR “mortality” OR “Viral Load”[Mesh] OR “Virus Physiological Phenomena”[Mesh] OR “Respiratory Distress Syndrome, Adult”[Mesh] OR “Respiratory Tract Infections”[Mesh] OR “Gastrointestinal Diseases”[Mesh] OR “Gastroenteritis”[Mesh] OR “Seizures”[Mesh] OR “Diarrhea”[Mesh] OR “Dehydration”[Mesh] OR “Water-Electrolyte Imbalance”[Mesh] OR “Kidney Failure, Chronic”[Mesh] OR “Shock”[Mesh] OR “Encephalomyelitis”[Mesh] OR “Mucocutaneous Lymph Node Syndrome”[Mesh] OR “Blood Coagulation Disorders”[Mesh] OR “Mortality”[Mesh]))

#### AND

(“vitamin C” OR “ascorbic acid” OR “Ascorbic Acid”[Mesh])

### Example search string in EMBASE

((coronavir* OR “coronavirus infections” OR covid* OR ncov* OR 2019-ncov OR 2019ncov OR “2019-novel CoV” OR HCoV* OR cov2 OR “cov 2” OR OC43 OR NL63 OR 229E OR HKU1 OR “sars coronavirus 2” OR “sars-like coronavirus” OR “Severe Acute Respiratory Syndrome” OR SARS OR sars-cov* OR sarscov* OR “Middle East Respiratory Syndrome” OR MERS OR MERS-CoV).ti,ab,kw. OR exp Coronavirus/ OR exp Coronaviridae infection/ OR exp severe acute respiratory syndrome/ OR exp SARS coronavirus/ OR exp Middle East respiratory syndrome coronavirus/)

#### AND

(((“adaptive immunity” OR “acquired immunity” OR “innate immunity” OR “cell-mediated immunity” OR “humoral immunity” OR “antibody formation” OR immunosuppression OR immunodepression OR “immunity impairment” OR “immune dysfunction” OR “lymphocyte function” OR “macrophage activity” OR “oxidative stress” OR “host defence” OR “immune response” OR inflammation OR “immune pathology” OR immunopathology OR “Macrophage activation syndrome” OR MAS OR “cytokine storm”).ti,ab,kw. OR exp immune system/ OR exp T lymphocyte/ OR exp inflammation/ OR exp immune deficiency/ OR exp oxidative stress/ OR exp macrophage activation/ OR exp cytokine storm/)

#### OR

((“viral load” OR “viral pathogen*” OR “viral replication” OR “viral mutation” OR “viral transmission” OR “acute respiratory distress syndrome” OR ARDS OR “hemophagocytic lymphohistiocytosis” OR HLH OR pneumonia OR bronchitis OR bronchiolitis OR “asthma exacerbation*” OR seizure* OR diarrhoea OR diarrhea OR “acute gastroenteritis” or dehydration or “electrolyte imbalance” OR “renal failure” OR “kidney failure” OR “multi-organ failure*” OR “multiple organ failure*” OR encephalomyelitis OR “Kawasaki disease” OR “Kawasaki syndrome” OR “Mucocutaneous Lymph Node Syndrome” OR coagulopathy OR death OR mortality).ti,ab,kw. OR exp virus load/ OR exp virus transmission/ OR exp virus shedding/ OR exp virus virulence/ OR exp virus characterization/ OR exp virus immunity/ OR exp virus infection/ OR exp virus cell interaction/ OR exp virus transcription/ OR exp virus inhibition/ OR exp respiratory tract infection/ OR exp gastrointestinal infection/ OR exp pneumonia/ OR exp virus pneumonia/ OR exp bronchitis/ OR exp bronchiolitis/ OR exp viral bronchiolitis/ OR exp asthma/ OR exp seizure/ OR exp diarrhea/ OR exp gastroenteritis/ OR exp viral gastroenteritis/ OR exp dehydration/ OR exp electrolyte disturbance/ OR exp kidney failure/ OR exp multiple organ failure/ OR exp encephalomyelitis/ OR exp mucocutaneous lymph node syndrome/ OR exp blood clotting disorder/ OR exp mortality/))

#### AND

((“vitamin C” OR “ascorbic acid”).ti,ab,kw. OR exp ascorbic acid/)

### Search terms for specific sections (used in concept 4)

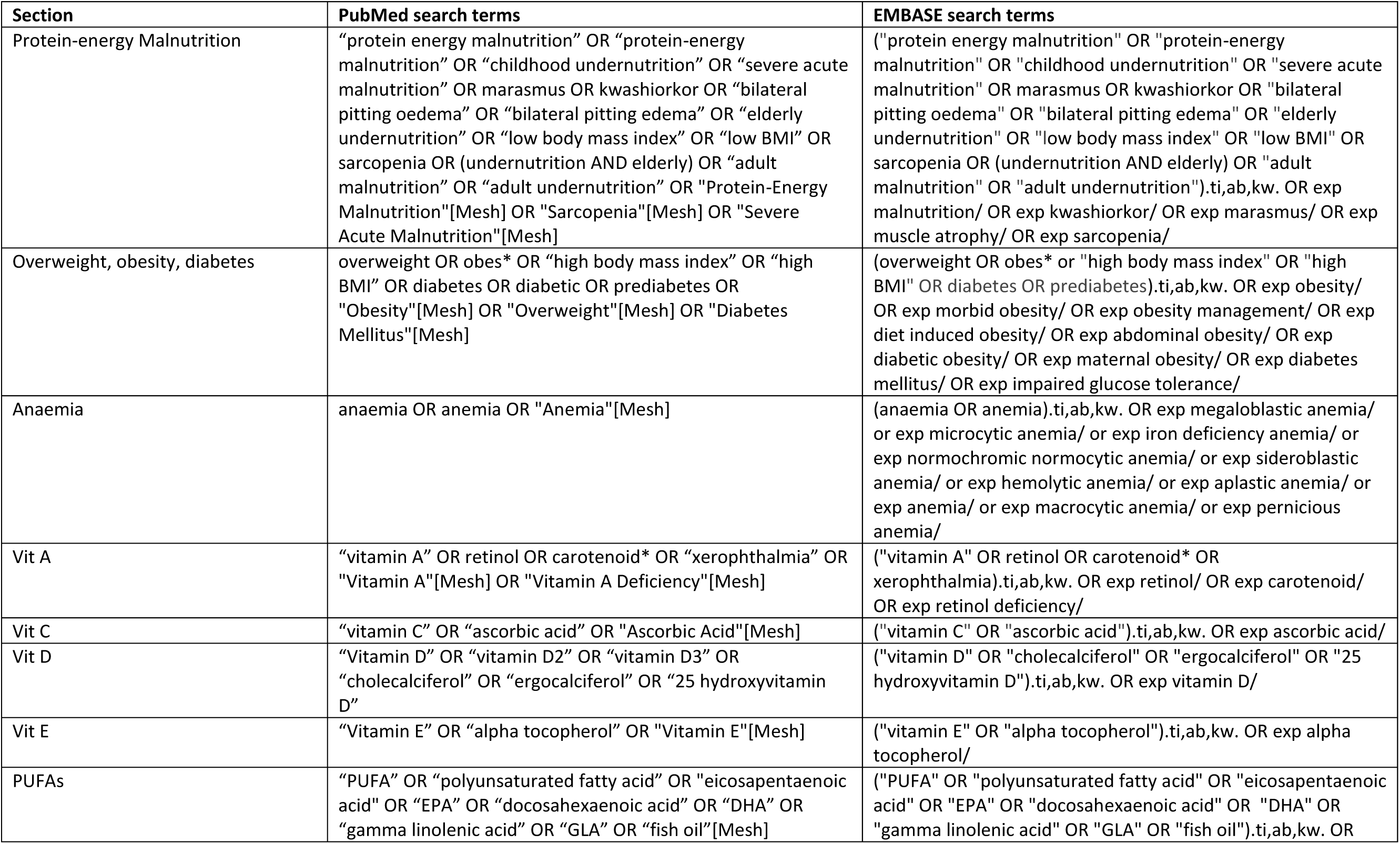
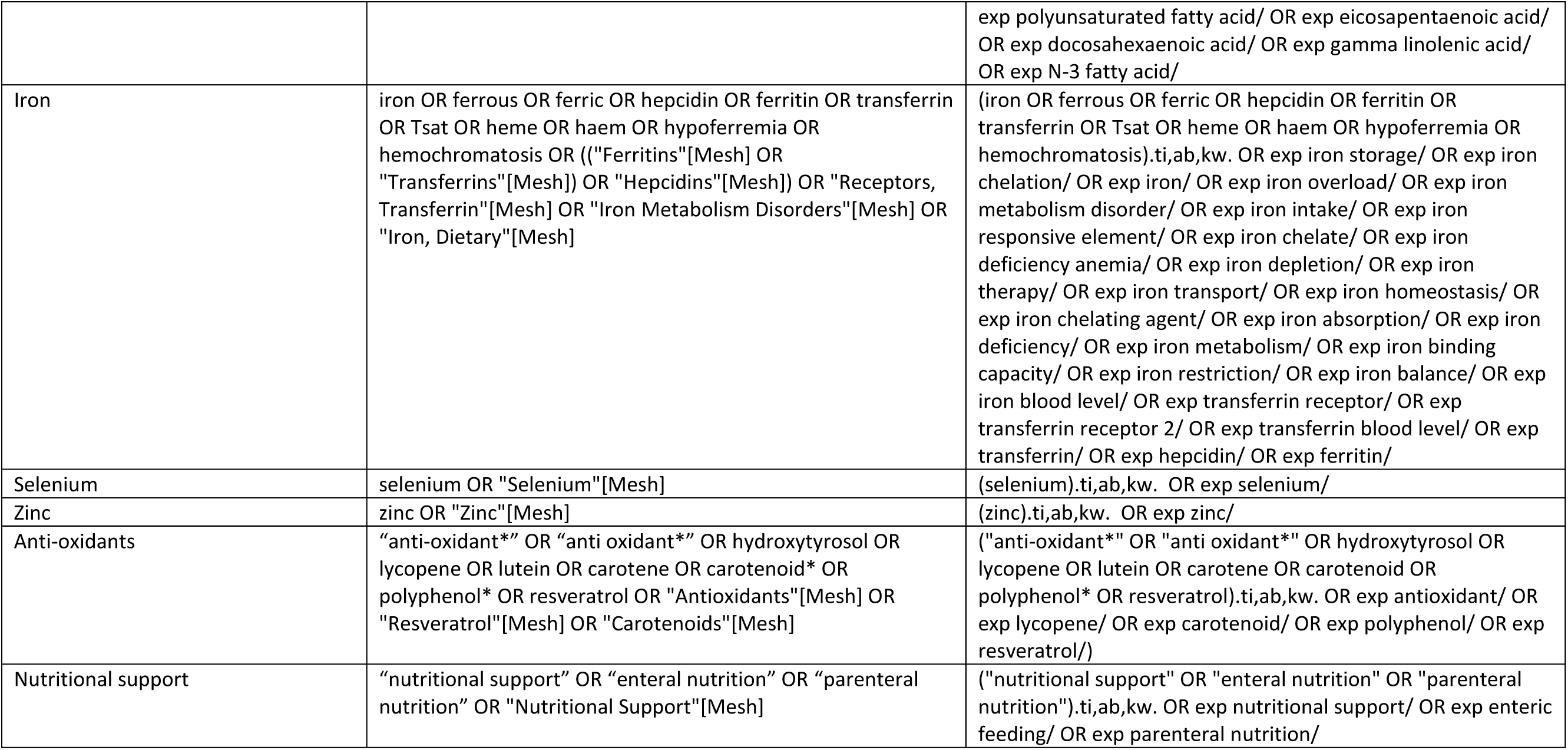

| Section | PubMed search terms | EMBASE search terms |
| --- | --- | --- |
| Protein-energy Malnutrition | "protein energy malnutrition" OR "protein-energy malnutrition" OR "childhood undernutrition" OR "severe acute malnutrition" OR marasmus OR kwashiorkor OR "bilateral pitting oedema" OR "bilateral pitting edema" OR "elderly undernutrition" OR "low body mass index" OR "low BMI" OR sarcopenia OR (undernutrition AND elderly) OR "adult malnutrition" OR "adult undernutrition" OR "Protein-Energy Malnutrition"[Mesh] OR "Sarcopenia"[Mesh] OR "Severe Acute Malnutrition"[Mesh] | ("protein energy malnutrition" OR "protein-energy malnutrition" OR "childhood undernutrition" OR "severe acute malnutrition" OR marasmus OR kwashiorkor OR "bilateral pitting oedema" OR "bilateral pitting edema" OR "elderly undernutrition" OR "low body mass index" OR "low BMI" OR sarcopenia OR (undernutrition AND elderly) OR "adult malnutrition" OR "adult undernutrition").ti,ab,kw. OR exp malnutrition/ OR exp kwashiorkor/ OR exp marasmus/ OR exp muscle atrophy/ OR exp sarcopenia/ |
| Overweight, obesity, diabetes | overweight OR obes* OR "high body mass index" OR "high BMI" OR diabetes OR diabetic OR prediabetes OR "Obesity"[Mesh] OR "Overweight"[Mesh] OR "Diabetes Mellitus"[Mesh] | (overweight OR obes* OR "high body mass index" OR "high BMI" OR diabetes OR prediabetes).ti,ab,kw. OR exp obesity/ OR exp morbid obesity/ OR exp obesity management/ OR exp diet induced obesity/ OR exp abdominal obesity/ OR exp diabetic obesity/ OR exp maternal obesity/ OR exp diabetes mellitus/ OR exp impaired glucose tolerance/ |
| Anaemia | anaemia OR anemia OR "Anemia"[Mesh] | (anaemia OR anemia).ti,ab,kw. OR exp megaloblastic anemia/ or exp microcytic anemia/ or exp iron deficiency anemia/ or exp normochromic normocytic anemia/ or exp sideroblastic anemia/ or exp hemolytic anemia/ or exp aplastic anemia/ or exp anemia/ or exp macrocytic anemia/ or exp pernicious anemia/ |
| Vit A | "vitamin A" OR retinol OR carotenoid* OR "xerophthalmia" OR "Vitamin A"[Mesh] OR "Vitamin A Deficiency"[Mesh] | ("vitamin A" OR retinol OR carotenoid* OR xerophthalmia).ti,ab,kw. OR exp retinol/ OR exp carotenoid/ OR exp retinol deficiency/ |
| Vit C | "vitamin C" OR "ascorbic acid" OR "Ascorbic Acid"[Mesh] | ("vitamin C" OR "ascorbic acid").ti,ab,kw. OR exp ascorbic acid/ |
| Vit D | "Vitamin D" OR "vitamin D2" OR "vitamin D3" OR "cholecalciferol" OR "ergocalciferol" OR "25 hydroxyvitamin D" | ("vitamin D" OR "cholecalciferol" OR "ergocalciferol" OR "25 hydroxyvitamin D").ti,ab,kw. OR exp vitamin D/ |
| Vit E | "Vitamin E" OR "alpha tocopherol" OR "Vitamin E"[Mesh] | ("vitamin E" OR "alpha tocopherol").ti,ab,kw. OR exp alpha tocopherol/ |
| PUFAs | "PUFA" OR "polyunsaturated fatty acid" OR "eicosapentaenoic acid" OR "EPA" OR "docosahexaenoic acid" OR "DHA" OR "gamma linolenic acid" OR "GLA" OR "fish oil"[Mesh] | ("PUFA" OR "polyunsaturated fatty acid" OR "eicosapentaenoic acid" OR "EPA" OR "docosahexaenoic acid" OR "DHA" OR "gamma linolenic acid" OR "GLA" OR "fish oil").ti,ab,kw. OR |
|  |  | exp polyunsaturated fatty acid/ OR exp eicosapentaenoic acid/<br>OR exp docosahexaenoic acid/ OR exp gamma linolenic acid/<br>OR exp N-3 fatty acid/ |
| Iron | iron OR ferrous OR ferric OR hepcidin OR ferritin OR transferrin<br>OR Tsat OR heme OR haem OR hypoferremia OR<br>hemochromatosis OR (("Ferritins"[Mesh] OR<br>"Transferrins"[Mesh]) OR "Hepcidins"[Mesh]) OR "Receptors,<br>Transferrin"[Mesh] OR "Iron Metabolism Disorders"[Mesh] OR<br>"Iron, Dietary"[Mesh] | (iron OR ferrous OR ferric OR hepcidin OR ferritin OR<br>transferrin OR Tsat OR heme OR haem OR hypoferremia OR<br>hemochromatosis).ti,ab,kw. OR exp iron storage/ OR exp iron<br>chelation/ OR exp iron/ OR exp iron overload/ OR exp iron<br>metabolism disorder/ OR exp iron intake/ OR exp iron<br>responsive element/ OR exp iron chelate/ OR exp iron<br>deficiency anemia/ OR exp iron depletion/ OR exp iron<br>therapy/ OR exp iron transport/ OR exp iron homeostasis/ OR<br>exp iron chelating agent/ OR exp iron absorption/ OR exp iron<br>deficiency/ OR exp iron metabolism/ OR exp iron binding<br>capacity/ OR exp iron restriction/ OR exp iron balance/ OR exp<br>iron blood level/ OR exp transferrin receptor/ OR exp<br>transferrin receptor 2/ OR exp transferrin blood level/ OR exp<br>transferrin/ OR exp hepcidin/ OR exp ferritin/ |
| Selenium | selenium OR "Selenium"[Mesh] | (selenium).ti,ab,kw. OR exp selenium/ |
| Zinc | zinc OR "Zinc"[Mesh] | (zinc).ti,ab,kw. OR exp zinc/ |
| Anti-oxidants | "anti-oxidant*" OR "anti oxidant*" OR hydroxytyrosol OR<br>lycopene OR lutein OR carotene OR carotenoid* OR<br>polyphenol* OR resveratrol OR "Antioxidants"[Mesh] OR<br>"Resveratrol"[Mesh] OR "Carotenoids"[Mesh] | ("anti-oxidant*" OR "anti oxidant*" OR hydroxytyrosol OR<br>lycopene OR lutein OR carotene OR carotenoid OR<br>polyphenol* OR resveratrol).ti,ab,kw. OR exp antioxidant/ OR<br>exp lycopene/ OR exp carotenoid/ OR exp polyphenol/ OR exp<br>resveratrol/) |
| Nutritional support | "nutritional support" OR "enteral nutrition" OR "parenteral<br>nutrition" OR "Nutritional Support"[Mesh] | ("nutritional support" OR "enteral nutrition" OR "parenteral<br>nutrition").ti,ab,kw. OR exp nutritional support/ OR exp enteric<br>feeding/ OR exp parenteral nutrition/ |

### Searches on clinical trial registries and pre-print servers are restricted to COVID-19 related studies, and use simplified search terms as below

| Section | Disease | Nutritional exposure |
| --- | --- | --- |
| Protein-energy Malnutrition | coronavirus AND | “Protein-energy malnutrition”<br>Undernutrition<br>Sarcopenia<br>“Severe acute malnutrition |
| Overweight, obesity, diabetes | coronavirus AND | Overweight<br>Obesity<br>Obese<br>Diabetes |
| Anemia | coronavirus AND | Anemia<br>Anaemia |
| Vit A | coronavirus AND | “Vitamin A”<br>Retinol<br>Carotenoid |
| Vit C | coronavirus AND | “Vitamin C”<br>“Ascorbic Acid” |
| Vit D | coronavirus AND | “Vitamin D”<br>Cholecalciferol<br>Ergocalciferol<br>“25 hydroxyvitamin D” |
| Vit E | coronavirus AND | “Vitamin E”<br>Tocopherol |
| PUFAs / anti-inflammatories | coronavirus AND | “Polyunsaturated fatty acids”<br>PUFA<br>Omega-3<br>Eicosapentaenoic Acid |
| Iron | coronavirus AND | Iron<br>Ferritin<br>Hepcidin<br>Transferrin |
| Selenium | coronavirus AND | Selenium |
| Zinc | coronavirus AND | Zinc |
| Anti-oxidants | coronavirus AND | Antioxidants<br>"Anti oxidants"<br>Anti-oxidants<br>"Free radical" |
| Nutritional support | coronavirus AND | "nutritional support"<br>"enteral nutrition"<br>"Parenteral nutrition" |

## Supplementary Material 3: Detailed Search Results per section

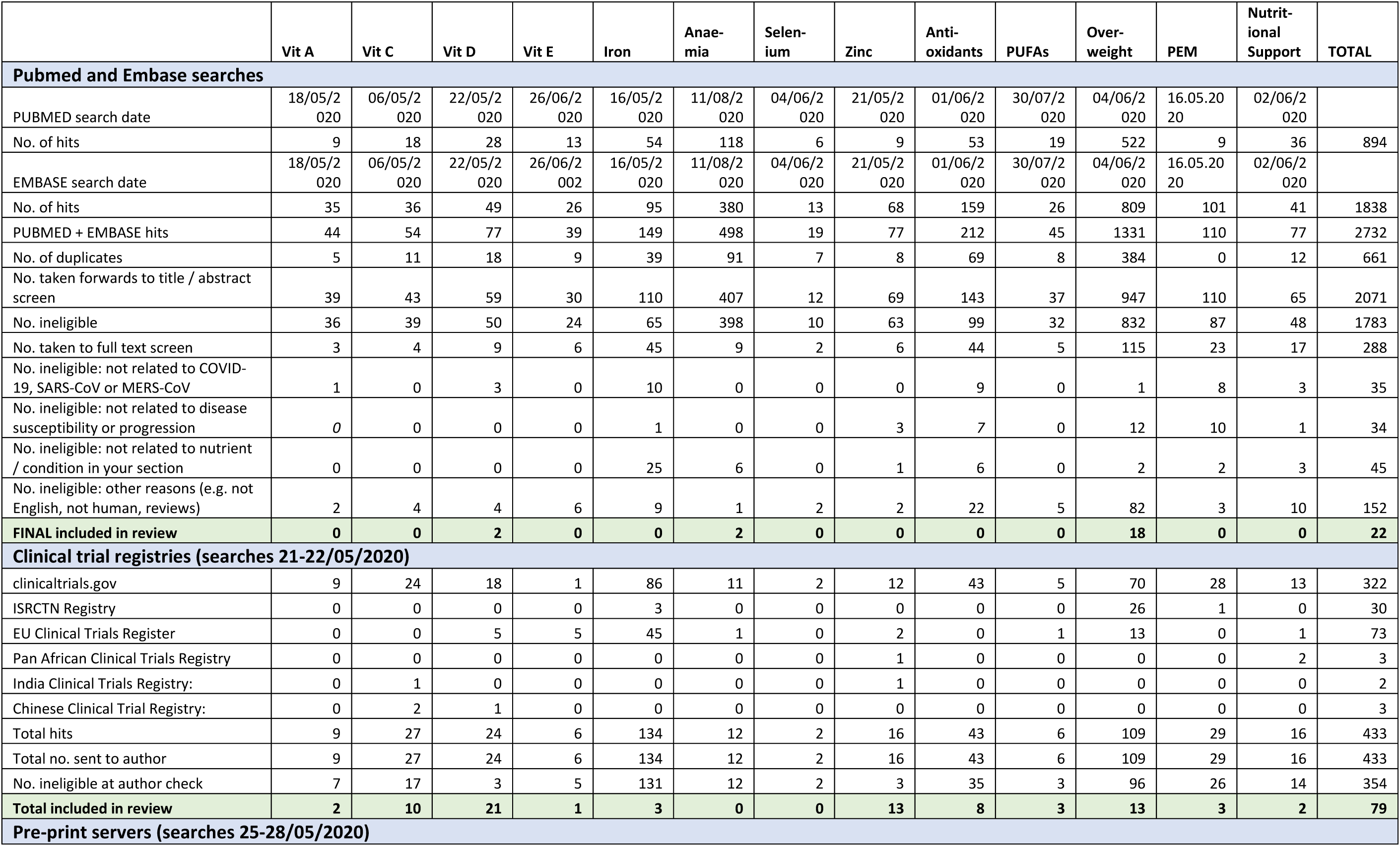
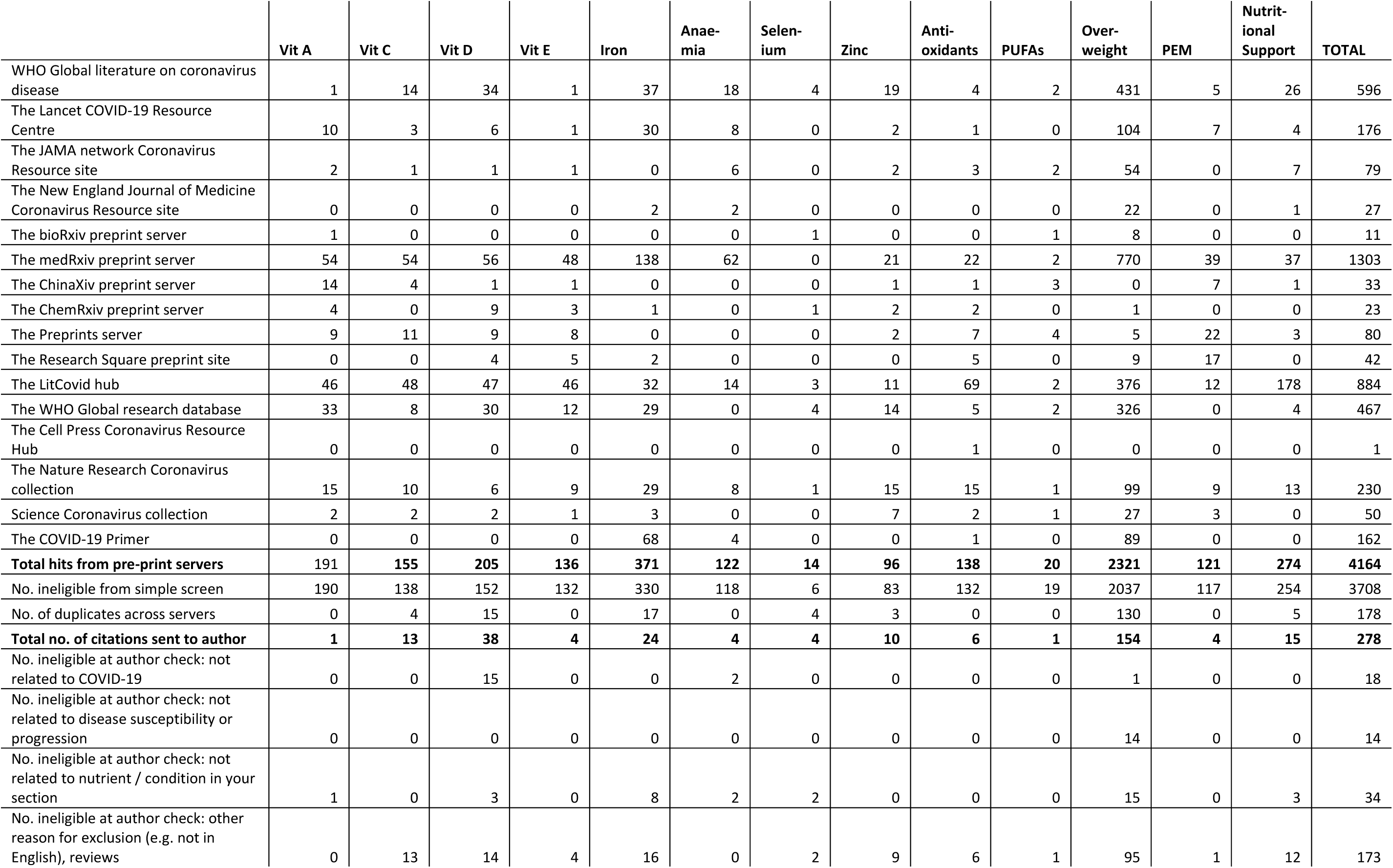
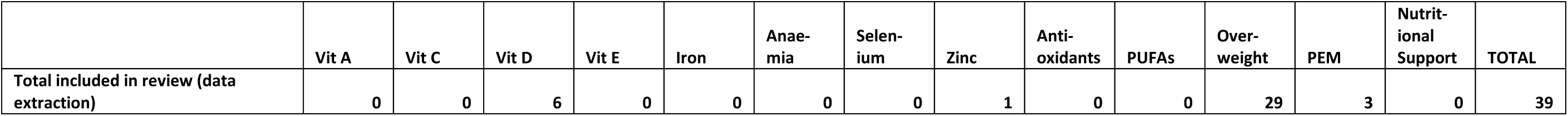

## Supplementary Material 4: Results from Clinical Trial Registries Search

| Section | Registry ID | Country | Trial design | Sample size | Population | Intervention | Comparator | Outcomes |
| --- | --- | --- | --- | --- | --- | --- | --- | --- |
| Vit A | NCT04323228 | Saudi Arabia | Double blind RCT | 30 | Hospitalised COVID-19 patients | 8fl oz Oral Nutrition supplement (ONS) enriched in eicosapentaenoic acid, gamma-linolenic acid and antioxidants (including 2840 IU vitamin A) | Isocaloric placebo | Serum ferritin level, cytokine storm parameters (interleukin-6, Tumor necrosis factor-α, and monocyte chemoattractant protein 1), C-reactive protein, total leukocyte count, differential lymphocytic count and neutrophil to lymphocyte ratio. |
| Vit A | NCT04360980 | Iran | Double blind RCT | 80 | Hospitalised no-ICU COVID-19 patients | Colchicine tablets -1.5 mg loading then 0.5 mg BID P.O | Standard of care including daily (vitamin C 3grams, 400 mg Thiamine, Selenium, Omega-3 500 mg daily, Vit A, Vit D, Azithromycin, Ceftriaxone, Kaletra 400 BID) | Primary outcomes: 1. CRPxN/R ratio change. 2. Clinical deterioration by the WHO definition including change in fever or O2 Saturation. 3. PCR Viral Load change in RT-PCR. 4. CT severity involvement index change in CT involvement. |
| Vit D | NCT04372017 | USA | Prospective, double-blind, randomized, placebo-controlled trial | 1739 | Healthcare workers and high-risk participants | Hydroxychloroquine - 800mg on day 1 followed by 400mg on days 2-5. | Vitamin D - IU1600 on day 1 and IU 800 on days 2-5 | COVID-19 status |
| Vit D | NCT04335084 | USA | Double-blind, randomized, placebo-controlled phase-IIa trial | 600 | Medical workers | Hydroxychloroquine, vit C, vit D, Zn | VitC, vitD, Zn | Prevention of COVID-19 symptoms |
| Vit D | EUCTR: 2020-001363-85 | Denmark | Randomized controlled trial | 206 | Nursing home residents >65y Not previously infected | 200 mg Hydroxychloroquine + unknown dose of vit D, Zn | No data | Prevention of COVID-19 symptoms |
| Vit D | EUCTR: 2020-002274-28 | Spain | non-blind, randomised, two-arm, no-control trial | 60 | Positive cases admitted to hospital > 18y | VitD (Videsil 100.000 UI) | None | Progression and clinical outcomes |
| Vit D | NCT04363840 | USA | non-blind, randomised, two-arm, no-control trial | 1080 | Newly diagnosed COVID-19 patients | Group 1 - Aspirin (81mg once daily for 14 days) & group 2 aspirin (81mg once daily for 14 days) + vitD (50,000 IU once weekly for 2 weeks) | Observation only | Hospitalization for COVID-19 |
| Vit D | NCT04360980 | Iran | Double-blind, randomized, intervention trial | 80 | Confirmed COVID-10 patients (Adults) | Colchicine (1.5 mg loading then 0.5 mg BID P.O) + std tx (vitamin C 3grams daily, 400 mg Thiamine, Selenium, Omega-3 500 mg daily, Vit A, Vit D, Azithromycin, Ceftriaxone, Kaletra 400 BID 10 days) | Standard tx (vitamin C 3grams daily, 400 mg Thiamine, Selenium, Omega-3 500 mg daily, Vit A, Vit D, Azithromycin, Ceftriaxone, Kaletra 400 BID 10 days) | CRP, clinical status, PCR viral load, CT severity involvement index |
| Vit D | NCT04334512 | USA | Randomized, Double-Blind, Placebo-Controlled Phase II intervention trial | 600 | Adults with diagnosis of COVID-19 | Hydroxychloroquine, azithromycin, vitC, vitD, Zn | VitC, vitD, Zn | Rate of recovery, reduction of symptomatic days |
| Vit D | NCT04399746 | Mexico | non-blind, non-randomised, control trial | 30 | Adults with confirmed COVID-19 (mild symptoms) | Ivermectin 6mg once daily in day 0,1,7 and 8; Azithromycin 500mg once daily for 4 days; Cholecalciferol 400 IU twice daily for 30 days | Observation only | Viral clearance, symptoms duration, O2 sat |
| Vit D | NCT04400890 | USA | Randomized Double-Blind Placebo-Controlled Trial | 200 | Adults 45y+ with mild COVID-19 symptoms | Resveratrol (unknown dose) + vitD (Vitamin D3 100,000 IU on day 1) | VitD (Vitamin D3 100,000 IU on day 1) | Hospitalization for COVID-19 |
| Vit D | NCT04344041 | France | Non-blinded Randomized Controlled Trial | 260 | Adults >=70y with COVID-19 | High dose vitD (400,000IU) | Low dose vitD (50,000IU) | Number of deaths |
| Vit D | NCT04366908 | Spain | Non-blinded Randomized Controlled Trial | 1008 | Adults 18 - 90 with PCR confirmed COVID-19 | VitD (Calcifediol: 524µg on day 1; 266 µg on days 3, 7, 14, 21 and 28) + best available tx (combination of drugs included in the current protocol of the Ministry of Health and/or complementary notes issued by the Spanish Agency of Medicines and Health Products (AEMPS)) | Best available tx (combination of drugs included in the current protocol of the Ministry of Health and/or complementary notes issued by the Spanish Agency of Medicines and Health Products (AEMPS)) | Admission to ICU, death |
| Vit D | NCT04351490 | France | Non-blinded Randomized Controlled Trial | 3140 | Institutionalized Adults 60y+ | Zinc: 15 mg x 2 per day during 2 months; 25-OH cholecalciferol (2000 IU) per day during 2 months | Usual tx | Survival rate |
| Vit D | NCT04385940 | Canada | Double-blinded, randomized, intervention trial | 64 | 17y+ with COVID-19 | High dose vitD (50,000IU) | Low dose vitD (1000IU) | Symptom recovery |
| Vit D | NCT04334005 | Spain | Double-blind, randomized, intervention trial | 200 | 40 - 70y with mild respiratory infection | Single dose of 25000 UI VitD + usual tx (NSAIDs, ACE2 inhibitor, ARB or thiazolidinediones) | Usual tx (NSAIDs, ACE2 inhibitor, ARB or thiazolidinediones) | Mortality |
| Vit D | EUCTR: 2020-001960-28 | Spain | Randomized Double-Blind Placebo-Controlled Trial | 108 | Hospital admission with positive COVID, 18y+ | 0,266 mg Calcifediol (unknown frequency) | Placebo | Progression and mortality |
| Vit D | NCT04395768 | Australia | Single-blind, randomized, control trial | 200 | 18y+ with active COVID-19 | Vit C (inpatients: Vitamin C (Sodium Ascorbate) 50mg/kg every 6hrs on day 1 followed by 100mg/kg every 6hrs (4x per day; 400mg/kg/day) for 7 days (average 28g/day; maximum dose of 50g/24hrs for those weighing more than 125kg) & outpatients: 200mg/kg x1 IV, then 1 gram PO three times per day for 7 days) PLUS standard treatment: Hydroxychloroquine 400mg (2x200mg) PO for 1 day, followed by 200mg PO per day for 6 days Azithromycin 500 mg PO on day 1 followed by 250 mg PO once daily for 4 days Zinc Citrate 30mg elemental zinc PO daily Vitamin D3 5,000iu PO daily for 14 days Vitamin B12 (Methylcobalamin) 500mcg PO daily for 14 days | Standard treatment: Hydroxychloroquine 400mg (2x200mg) PO for 1 day, followed by 200mg PO per day for 6 days Azithromycin 500 mg PO on day 1 followed by 250 mg PO once daily for 4 days Zinc Citrate 30mg elemental zinc PO daily Vitamin D3 5,000iu PO daily for 14 days Vitamin B12 (Methylcobalamin) 500mcg PO daily for 14 days | Symptoms, duration of hospital stay, ventilation, mortality |
| Vit D | NCT04386850 | Iran | multicentre randomized double-blinded placebo-controlled clinical trial | 1500 | 18-75y (1) diagnosed (2) prevention | 25 mcg of 25(OH)D3 once daily - for 2 months | Placebo for 2 months | Infection, severity, hospitalization, disease duration, death, O2 support |
| Vit D | NCT04394390 | Turkey | prospective cohort study (2 months follow-up) | 100 | Children and adults | n/a - measure "exposure" is Vitamin D levels in positive COVID-19 patients | n/a | Disease severity |
| Vit D | NCT04386044 | UK | cross-sectional (hospitalised patients) & prospective cohort study (6 months follow-up of participants recruited from GP practice) | 1800 | Adults, infected and uninfected | n/a - measured "exposure" is Vitamin D levels in positive COVID-19 patients & Vitamin D levels in GP patients | n/a | Cross sectional: Death, O2 therapy - Cohort: Infection with COVID 19 |
| Vit D | NCT04370808 | Portugal | Prospective cohort study | 500 | 18y+ with active COVID-19 | n/a - measured "exposure" is Vitamin D levels / genetic variants in vitamin D-related genes | n/a | COVID 19 severity and death |
| Vit D | ChiCTR2000031163 | China | Prospective cohort study | 80 | 0.1 - 85 y males | n/a - Measured "exposure" is Vitamin D deficiency | n/a | Progression, treatment outcome and prognosis of COVID-19 |
| Iron | NCT04389801 | Egypt | Single-blind, placebo-controlled RCT | 200 | Hospitalised COVID-19 with chest tightness | Deferoxamine (desferal), Initial 1000 mg at 15 mg.kg.hr (1g) Subsequent 500mg at 125 mg.hr (0.5g) | 5% glucose | 14d mortality |
| Iron | NCT04361032 | Tunisia | Open-label, Multicentric, Comparative, Randomized Study | 260 | ARDS ICU COVID-19, (18-80 years) | Deferoxamine (desferal), 40 mg.kg.d x 14d (2.8g) | Tocilizumab, 8mg.kg.d | 90d mortality |
| Iron | NCT04333550 | Iran | Double-blind, RCT | 50 | Mild, moderate or severe pneumonia | Deferoxamine (desferal), dose not stated | Standard treatment | 20d mortality |
|  |  |  |  |  | COVID-19, (3-99 years) |  |  |  |
| Zinc | EudraCT Number: 2020-001449-38<br>ClinicalTrials.gov Identifier: NCT04373733 | UK | Randomised non-blinded randomised trial | 450 participants | London > 18 y COVID patients | **Intervention 1**<br>(Favipiravir & Standard of Care) Favipiravir: Day 1 1800mg twice per day, Days 2-10 800mg twice per day **Intervention 2**<br>(Hydroxychloroquine, azithromycin, zinc & Standard of Care):<br>Hydroxychloroquine: Day 1 400mg twice per day, Days 2-10 200mg twice per day; Azithromycin: Day 1-3 250mg once per day; Zinc-sulphate: Days 1-10 125mg twice per day | Standard of care | Outcome: Time to clinical improvement |
| Zinc | NCT04377646 | Tunisia | Double blind randomised placebo-controlled trial with 3 arms | 660 | Tunisia; 18 - 65 y, COVID negative | **Active comparator:** HCQ & zinc:<br>Hydroxychloroquine 400 mg at day 1 and day 2, then a weekly dose of 400 mg up to 2 months. Zinc 15 mg at daily dose up to 2 months | Double placebo of HCQ and zinc | Primary outcome: Frequency of confirmed SARS CoV2 infection |
| Zinc | NCT04335084 | USA | A Randomized, Double-Blind, Placebo-Controlled Phase IIa Study | 600 | USA; Medical workers who are exposed to COVID-19 and as such are at higher risk of infection | NB No information on dosing levels.<br>**Intervention:**<br>Hydroxychloroquine, Vitamin C, Vitamin D, Zinc. | Vitamin C, Vitamin D, Zinc | Primary outcome: Prevention of COVID symptoms over 24 weeks |
| Zinc | NCT04334512 | USA | Randomized, Double-Blind, Placebo-Controlled Phase II interventional Study | 600 | USA; 18 y + with COVID diagnosis | NB No information on dosing levels.<br>**Intervention:** HCQ, Azithromycin, vit C, vit D, zinc | Vit C, vit D, zinc. | Primary outcome: Rate of recovery of mild or moderate COVID-19 |
| Zinc | NCT04323228 | Saudi Arabia; 18-65y | Randomized, Double-Blind, Placebo-Controlled trial | 30 | Saudi Arabia; 18-65y; confirmed COVID-19 but stable condition | **Intervention:** will receive daily oral nutrition supplement (ONS) enriched in eicosapentaenoic acid, gamma-linolenic acid and antioxidants. The composition of one can (8 fl oz) of the intervention-ONS includes: 14.8 g protein, 22.2 g fat, 25 g carbohydrate, 355 kcal, 1.1 g EPA, 450 mg DHA, 950 mg GLA, 2840 IU vitamin A as 1.2 mg β-carotene, 205 mg Vitamin C, 75 IU vitamin E, 18 ug Selenium, and 5.7 mg Zinc. | Iso-caloric - isonitrogenous product (by the same manufacture) and served in cans with the same colour and shape. | Primary outcome: Nutrition risk screening and biochemical markers |
| Zinc | NCT04342728 | USA | Randomised open label trial | 520 | USA; > 18 y ; outpatients who test positive for COVID-19 | **Ascorbic acid:** 8000 mg of ascorbic acid divided into 2-3 doses/day with food.<br>**Zinc gluconate:** 50 mg of zinc gluconate to be taken daily at bedtime<br>**Ascorbic acid & zinc gluconate:** 8000 mg of ascorbic acid divided into 2-3 doses/day with food and 50 mg of zinc gluconate to be taken daily at bedtime. | Standard of care. | Primary outcome: symptom reduction over 28 days |
| Zinc | NCT04384458 | Brazil | Open non-blinded randomised trial | 400 | Brazil; healthcare workers aged 18 to 70 y. No COVID | **Active Comparator:**<br>Hydroxychloroquine Oral hydroxychloroquine 400 mg twice a day on day 1, one 400 mg tablet on day 2, 3, 4, and 5, followed by one 400 mg tablets every 05 days until day 50th associated with 66 mg of zinc sulphate. | No intervention (Control) Subjects who don't want not to receive the study drug, but agree to participate by signing the informed consent, will form the control group | Primary outcome: proportion of participants in whom there was a clinical finding of COVID-19 or number of symptomatic COVID-19 infections |
| Zinc | EudraCT Number: 2020-001363-85 | Denmark | Randomised controlled Open trial | 206 Nursing home residents > 65 y | Demark | **Intervention:**<br>Hydroxychloroquine 200mg, vit d & zinc (dose not found) |  | Outcome: severity of the disease, hospitalization rate, and death in nursing home residents. |
| Zinc | CTRI/2020/05/025215 | India | Randomized, parallel Group Trial | 50 per group | India; COVID patients aged 18 - 55 y | **Intervention 1.** Kabasura Kudineer 60 ml bd for 14 days | Vitamin C - 60000IU OD for 14 days Zinc supplementation - 100mg od for 14 days | Outcome: Reduction in incidence of clinical symptoms of COVID |
| Zinc | NCT04370782 | USA | Randomised, non-blinded two arm (no control) clinical trial | 750 | USA; > 30 y; high initial clinical suspicion of COVID-19 | **<u>Experimental Arm 1:</u>**<br>**<u>Hydroxychloroquine, Azithromycin & zinc sulphate</u>**<br>(Hydroxychloroquine 400mg twice a day (BID) on day 1, followed by 200mg BID for days 2-5; Azithromycin 500mg on day 1, followed by 250mg once daily for days 2-5, zinc sulphate 220mg once daily for 5 days).<br>**<u>Experimental Arm 2:</u>**<br>**<u>Hydroxychloroquine, Doxycycline & zinc sulphate</u>**<br>(Hydroxychloroquine 400mg twice a day (BID) on day 1, followed by 200mg BID for days 2-5; Doxycycline 200mg once daily for days 2-5, zinc sulphate 220mg once daily for 5 days) | NA | Primary outcomes: 1. Time to resolution of symptoms (day 5, 14 and 21), 2. number of participants hospitalized, 3. ICU Length of stay. 4. Ventilator time frame |
| Zinc | PACTR202005622389003 | Senegal | Randomised open label-controlled trial | 384 / 128 per group | Senegal; Patients > 18 < 65 y with confirmed COVID | **<u>Intervention 1:</u>**<br>Hydroxychloroquine will be given on 3 times daily (200mg - 200 mg - 200 mg) during 6 days<br>Azithromycin will be administered on a single daily dose: 500 mg the first day followed by 250 mg from day2 to day 5 - Intervention <u>2</u>.<br>Hydroxychloroquine will be given on 2 times daily | 3. Control: Zinc (20mg / d) | Death at day 7, time to first negative PCR after treatment initiation. Biochemical parameters. Haematological parameters, ECG abnormalities |
|  |  |  |  |  |  | (200mg - 200 mg) during 6 days Azithromycin will be administered on a single daily dose: 500 mg the first day followed by 250 mg from day2 to day 5 |  |  |
| Zinc | NCT04351490 | France | Randomised open label | 3140 | France; 60 y plus | **Experimental:** Zinc gluconate capsule 15mg x 2 per day during 2 months + 25-OH cholecalciferol (2000 IU) per day during 2 months | No intervention: Usual treatment | Primary outcome: Survival rate in asymptomatic subjects at inclusion |
| Zinc | NCT04395768 | Australia | Multi-centre, International, single - blinded Randomized Trial: | 200 participants | Australia > 18 y active diagnosis | **Experimental:** Vitamin C Participants will receive vitamin C in addition to active comparator treatment: **Inpatients:** IV Vitamin C (Sodium Ascorbate) 50mg/kg every 6hrs on day 1 followed by 100mg/kg every 6hrs (4x per day; 400mg/kg/day) for 7 days (average 28g/day; maximum dose of 50g/24hrs for those weighing more than 125kg). Can be converted to 1 gram three times per day PO on hospital discharge) **Outpatients:** Vitamin C Outpatient trial: 200mg/kg x1 IV, then 1-gram PO three times per day for 7 days | Hydroxychloroquine 400mg PO twice a day for 1 day, followed by 200mg PO two times a day for 6 days Azithromycin 500 mg PO on day 1 followed by 250 mg PO once daily for 4 days Zinc Citrate 30mg elemental zinc PO daily Vitamin D3 5,000iu PO daily for 14 days Vitamin B12 (Methylcobalamin) 500mcg PO daily for 14 days | Primary outcomes: 1. Symptoms 2. Length of hospital stay 3. Invasive mechanical ventilation |
| Nutritional support | NCT04365816 | France | Prospective cohort | 403 | Patients discharged from hospital after admission with covid-19 | N/A | N/A | Food intake at 1-month post discharge; weight variation during the infection; factors limiting food intake; implemented nutritional strategy; pre-existing chronic disorders; covid-19 repercussions |
| Nutritional support | NCT04274322 | China | Prospective cohort | 117 | ICU patients | N/A | N/A | Validate the use of NUTRIC score nutritional risk assessment tool in Chinese ICU patients with covid-19; 28-day all-cause mortality; all cause infection; rate of complications; length of ICU stay; duration of mechanical ventilation. |
| Vitamin C | NCT03680274 | Canada | Double-blind RCT | 800 | Septic ICU (inc COVID) | High dose intravenous vitamin C (HDIVC) 200 mg.kg.d x 4d (56g) | Dextrose 5% in water (D5W) or normal saline (0.9% NaCl). | 28-day mortality. |
| Vitamin C | NCT04264533 | China | Double-blind RCT | 140 | Severe viral pneumonia | HDIVC 340 mg.kg.d x 7days (168g) | 50ml water for injection | 28-day ventilator free days |
| Vitamin C | NCT04323514 | Italy | Open label non-randomized trial | 500 | COVID-19 pneumonia | Diet + IVC 140 mg.kg.d x 1d (10g) | N/A (Single group assignment). | 3-day mortality |
| Vitamin C | NCT04344184 | USA | Double-blind RCT | 200 | COVID-19 pneumonia | HDIVC 300 mg.kg.d x 3d (63g) | Dextrose 5% Water | 28-day ventilator free days |
| Vitamin C | NCT04357782 | USA | Open label non-randomized trial | 20 | Mild/sev deoxygenation | HDIVC 200 mg.kg.d x 4d (56g) | N/A (Single group assignment). | Incidence of adverse events |
| Vitamin C | NCT04363216 | USA | Open label randomized trial | 66 | Hospitalised COVID-19 | 300/600/900 mg.kg.d x 6d (126/252/378g) | Routine care | 3 day 'clinical improvement (50% reduction in the highest flow rate of oxygen during the 72 hour period OR 50% reduction in the most frequent use of bronchodilators within a 12-hour window within the 72-hour period OR hospital discharge). |
| Vitamin C | NCT04395768 | Australia | Double-blind RCT | 200 | 'COVID-19' | 200 mg.kg.d x 1d + 400 mg.kg.d x 7d (210g) | No vitamin C | 15 day symptoms since enrolment. |
| Vitamin C | NCT04401150 | Canada | Double-blind RCT | 800 | Hospitalised COVID-19 | HDIVC 200 mg.kg.d x 4d (56g) | Normal saline (0.9% NaCl) or dextrose 5% in water (D5W) in a volume to match the vitamin C. | 28 day mortality or persistent organ dysfunction (POD). |
| Vitamin C | ChiCTR 2000032716 | China | RCT | 12 | Severe/critical COVID-19 pneumonia | HDIVC Dose Not stated | Not stated | C-Reactive protein, lymphocytes, CD4+ |
| Vitamin C | ChiCTR 2000032717 | China | RCT | 60 | Mild/severe COVID | Bolus IVC 166 mg.kg.d x 1d | Not stated | Recovery time |
| Poly-unsaturated fatty acids (PUFAs) and antioxidants | NCT04323228 | Saudi Arabia | Double-blind RCT | 30 | Hospitalised COVID-19 | Oral eicosapentaenoic acid, gamma-linolenic acid and antioxidants. 1.1g eicosapentaenoic acid (EPA), 450 mg docosahexaenoic acid (DHA), 950 mg gamma linolenic acid (GLA), 2840 IU vitamin A as 1.2 mg β-carotene, 205 mg Vitamin C, 75 IU vitamin E, 18 ug Selenium, 5.7 mg Zinc | Placebo | 3 month change in score of Nutrition risk screening-2002 (NRS-2002) at end. |
| Poly-unsaturated fatty acids (PUFAs) | NCT04335032 | USA | Open label non-randomized trial | 240 | Hospitalised COVID-19 | Eicosapentaenoic acid gastro-resistant capsules. 2g daily eicosapentaenoic acid free fatty acid. | N/A (Single group assignment). | Time to treatment failure within 28d |
| Poly-unsaturated fatty acids (PUFAs) | NCT04460651 | Latin America | Double-blind RCT | 1500 | Healthcare providers at risk of COVID | Icosapent ethyl (IPE). 8g IPE days 1-3<br>4g IPE days 4-60 | Placebo | PCR or IgG positive for SARS-CoV-2 by day 60. |
| Overweight/obesity/diabetes | NCT04396106 | USA | Double-blind RCT | 190 | Moderate COVID patients with history of obesity (BMI>30), diabetes and hypertension age 45 to 80years | AT-527 at 550mg tablets on day 1 and twice everyday for 5 days. | Placebo | 14 day proportions (active vs. placebo) of subjects with progressive respiratory insufficiency. |
| Overweight/obesity/diabetes | NCT04391738 | France | Retrospective cohort | 1200 | Patients admitted to Intensive Care Unit with SARS-CoV-2 | N/A | N/A | 3 month relationship between body mass index (BMI) and SARS-CoV-2 |
| Overweight/obesity/diabetes | NCT04391686 | France | Observational Cohort | 90 | COVID-19 patients admitted at ICU with BMI>30 | N/A | N/A | The resting energy expenditure (in Kcal / 24h) measured by indirect calorimetry during the stay in intensive care (3 months later). |
| Overweight/obesity/diabetes | NCT04390555 | Switzerland | Observational Cohort | 1500 | Hospitalized COVID-19 patients with pre-existing cardiovascular diseases and/or cardiovascular risk factors (diabetes mellitus, arterial hypertension and/or dyslipidaemia ) | N/A | Patients with COVID-19 without pre-existing cardiac involvement. | In-hospital mortality, 30 days. |
| Overweight/obesity/diabetes | NCT04390074 | Sweden | Case-control | 9905 | ICU patients | N/A | Age- and sex-matched controls are drawn from all residents of Sweden by Statistics Sweden. | Odds of intensive care treated patients with COVID-19 having been diagnosed with diabetes type I, diabetes type II, obesity and other co-morbidities (6 months). |
| Overweight/obesity/diabetes | NCT04384471 | Canada | Cross-sectional | 384 | People living with type 1 diabetes in Quebec and are registered to a registry. | N/A | N/A | Self-reported acute-diabetes complication. Severe hypoglycaemia and diabetic ketoacidosis |
| Overweight/obesity/diabetes | NCT04382794 | Italy | Case-control | 338 | The anonymous data of diabetic patients hospitalized for COVID19 in the hospitals participating in the study will be collected | DMT2 COVID19 positive patients treated with Sitagliptin. | DMT2 COVID19 positive patients not treated with Sitagliptin | Clinical parameter of acute lung disease |
| Overweight/obesity/diabetes | NCT04371978 | Israel | Open label randomized trial | 100 | Hospitalised COVID-19 patients with type 2 diabetes. | Linagliptin 5 mg PO once daily | Standard of care | Time to clinical change (within 28 days) |
| Overweight/obesity/diabetes | NCT04365634 | China | Case-control | 306 | Hospitalised COVID-19 patients with and without diabetes. | N/A | N/A | The predictive factors associated with hospitalized death of patients with COVID-19 (28 days). |
| Overweight/obesity/diabetes | NCT04365517 | Italy | Open label randomized trial | 170 | Patients hospitalized for COVID-19 and suffering from type 2 diabetes. | Sitagliptin at an adjusted dosage for estimated glomerular filtrate: 100 mg once daily (estimated glomerular filtration rate less than or equal to 45 mL / min / 1.73 m2 ) or 50 mg (estimated glomerular filtration rate 30-45 mL / min / 1.73 m2) in combination or not with insulin | Standard of care | Time to clinical improvement within 1 month. Clinical parameter of acute lung disease (1 month). |
| Overweight/obesity/diabetes | NCT04341935 | USA | Open label randomized trial | 20 | Hospitalized for COVID-19 with type 2 diabetes. | 5 mg Linagliptin administered by mouth once daily | Standard of care | Changes in glucose levels up to 2 weeks. |
| Overweight/obesity/diabetes | NCT04324736 | France | Retrospective cohort | 5497 | Diabetic patients treated for COVID-19 in a hospital centre and non-diabetic patients treated for COVID-19 | N/A | N/A | Assess the prevalence of severe forms among hospitalized patients with diabetes and COVID-19 within 1 month. |
| Overweight/obesity/diabetes | NCT04324684 | Italy | Case-control | 198 | Subjects hospitalized for COVID-19 pneumonia presenting with complications including diabetes and obesity. | N/A | N/A | Time to improvement within 3 weeks. |
| Antioxidants | NCT04400890 | USA | Double-blind RCT | 200 | Mild COVID-19 with symptoms <7 days | Resveratrol QID x 15 days | Placebo | Hospitalisation 21 days from randomization. |
| Antioxidants | NCT04394208 | Egypt | Double-blind RCT | 50 | Patients with COVID-19 pneumonia | 420 mg/day Silymarin in 3 divided doses | Placebo | Time to clinical improvement (7-28 days) |
| Antioxidants | NCT04382040 | Israel | Double-blind RCT | 50 | COVID-19+; in stable to moderate condition (not requiring ICU admission) | 6 mg Artemisinin, 20 mg Curcumin, 15 mg Frankincense and 60 mg vitamin C given daily in two divided doses, on Days 1 and 2. | Placebo | Time to clinical improvement, percent of patients with adverse events (24 hours). |
| Antioxidants | NCT04377789 | Turkey | Non-randomised | 50 | At moderate-high risk for COVID19 (prophylaxis group), COVID19+ (treatment group) | 500 (prophylaxis group) or 1000mg (treatment group) Quercetin. Time of treatment unclear. | Placebo | Prevalence of COVID-19 (prophylaxis group). Mortality rate (treatment group) |
| Antioxidants | NCT04374461 | USA | Non-randomised | 86 | COVID-19+, Arm A: Admission to an ICU; Arm B: Hospital admission but not requiring mechanical ventilation or admission to an ICU | N-acetylcysteine IV 6 g/day | None | Arm A: number of patients who are successfully extubated and/or transferred out of critical care due to clinical improvement Arm B: number of patients who are discharged from the hospital due to clinical improvement |
| Antioxidants | NCT04353128 | Spain | Double blind RCT | 450 | COVID-19 negative healthcare workers | Melatonin 2 mg/day for 12 weeks | Placebo | SARS-CoV-2 infection rate up to 12 weeks. |
| Antioxidants | NCT04370288 | Iran | Single blind RCT | 20 | COVID-19 hospitalised patients | Methylene Blue (1 mg/kg) along with vitamin C (1500 mg/kg) and N-acetyl Cysteine (1500 mg/kg) orally or intravenously | Standard of care | Proportion of patients remaining free of need for mechanical ventilation |
| PEM | NCT04386460 | France |  | 100 | Nice University Hospital, France- adult patients attending dental clinic and referred to their Physician for assessment and nutritional Care | N/A | N/A | Body Mass Index evolution from baseline at 1 and 3 months |
| PEM | NCT04350073 | USA |  | 120 | US, Duke University ICU, Group 1: COVID-10 patients with respiratory failure admitted to the ICU, Group 2 (controls) Non-COVID-19 respiratory failure patients requiring mechanical ventilation > 48 h receiving similar ICU standards of care at Duke | Q-NRG Metabolic Cart Device; MuscleSound Ultrasound Multifrequency Bioimpedance Spectroscopy | Standard of care | Metabolic and nutritional needs of COVID-19 Patients; Cardiac Output and Cardiac Measures (non-invasive) in COVID-19 patients |
| PEM | NCT04346212 | Spain |  | 100 | Hospital de Mataró, Spain. Patients infected by SARS-CoV-2 at the Hospital de Mataró, Hospital de St. Jaume i Sta. Magdalena and other medicalized facilities in Mataró. | N/A | N/A | Prevalence of oropharyngeal dysphagia according to a clinical assessment tool, the Volume-Viscosity Swallowing Test (V-VST). |

## Notes

### Competing Interest Statement

The authors have declared no competing interest.

### Author Declarations

No ethical approval was sought for this systematic review of publicly available secondary data.

### Summary of Updates

Author 2 spelling of first name corrected.

